# Prevalence of HIV and Syphilis and their Co-infection among Men Having Sex with Men in Asia: A Systematic Review and Meta-analysis

**DOI:** 10.1101/2021.12.21.21268191

**Authors:** Sultan Mahmud, Md Mohsin, Abdul Muyeed, Sorif Hossain, Md Mynul Islam, Ariful Islam

## Abstract

**Background:** Studies found that the group of men who have sex with men (MSM) is at a very high level of risk of HIV and sexually transmitted infections (STIs) in Asian regions due to multiple reasons. Although the prevalence of HIV among general people in Asia is considered low, the prevalence of HIV and Syphilis among MSM in this region was found very high and usually, it goes unnoticed. This study aimed to inspect the prevalence and trends of HIV, Syphilis, and their co-infection among MSM in Asia.

**Methods:** A systematic search was performed on January 5, 2021, in PubMed, MEDLINE, and Google Scholar databases. To evaluate the heterogeneity, Q-tests and were used. To explore the publication bias, Eggers’ test and funnel plot were used. The random-effect model and subgroup analysis were performed due to the significant heterogeneity.

**Results:** A total of 2,872 articles were identified, and 66 articles of high quality were included in the final analysis. The overall prevalence of HIV among MSM and Syphilis was estimated considering 69 estimates from 66 studies whereas 19 estimates of co-infection were found in 17 studies. The pooled HIV prevalence was 8.48% (CI: 7.01-9.95) and the pooled Syphilis prevalence was 9.86% (CI: 8.30-11.41) with significant heterogeneity and publication bias. The pooled prevalence of HIV and Syphilis co-infection was 2.99% (CI: 1.70-4.27) with significant heterogeneity and no publication bias. The HIV, Syphilis, and HIV and Syphilis co-infection prevalence estimates exhibited an upward trend during 2002-2017.

**Conclusions:** HIV, Syphilis, and their co-infection are quite prevalent among MSM in the Asia-Pacific region. Integrated and intensified intervention strategies, HIV testing, and improved access to antiretroviral treatment as well as increased awareness are needed to reduce HIV, Syphilis, and their co-infection among the discussed vulnerable group.

## 1 Introduction

People who are sexual minorities and marginalized and disproportionately affected by HIV— particularly, homosexual men, bisexual men, and male-to-female transgender people, are known together as ‘‘men who have sex with men (MSM)’’ [1]. The MSM is the most vulnerable group of people who are being at a high level of risk of HIV and sexually transmitted infections (STIs) in Asia due to unprotected anal sex, heterosexuality, multiple sex partners, etc [2-5]. Globally in 2018, the chance of being infected with HIV in gay men was 22 times higher than the all-adult men [6]. The estimate of new infection rates among MSM or gay men globally is 17% and in Asia and the Pacific region, it is 30%, which is the second-highest after the Latin America region even though, most countries in Asia maintain very low (<0.1 %) to low (<1%) HIV prevalence in the general population [1]. A high prevalence has been reported among MSM (between 10% to 30%) in India [7], Thailand [8, 9], China [10-14], Cambodia [15], Indonesia, and Myanmar [16]. The HIV infection among MSM is still increasing [17-19] and it has become a threat in those Asian countries [20]. The reasons behind this might be poor HIV prevention programs, HIV testing services, as well as sexual health services in this region [19]. It is assumed that the behavior of this specific group of people is strongly correlated to HIV infection and sexually transmitted infection (STI) risk. Therefore, “MSM” has been used as an epidemiologic term to recognize men based on behavior rather than identity [21, 22]. In most of the countries in Asia where they are facing stigma, discrimination, and criminal sanctions, they are also known as “hidden” or “hard to find” populations for HIV prevention care and treatment and HIV surveillance [23-25]. Those stigmas, discrimination, criminal sanctions, and sociocultural heteronormative pressures influence them to have sex with women although they have diverse socio-cultural and behavioral contexts of sexuality [2, 22]. Due to the hidden characteristics, low-risk perception, unwillingness to disclose attitudes, and scarcity of community communications of MSMW, they may not be sufficiently reached by sexual health and HIV prevention program [26].

Syphilis is a sexually transmitted disease that is also acquainted with syphilitic ulcers to shatter the mucosal barriers. Syphilis increases the likelihood of HIV infection. A certain epidemic of Syphilis occurred in the nearest past among MSM in different parts of the world [27]. An outbreak of Syphilis among MSM has been observed in the 2000s in many developed countries [28-30]. Among the HIV-positive MSMs, the Syphilis incidence rate has been observed 2.9 to 6.2 per hundred people in the Western world [31-33]. Even, in some cities of Asian countries, the Syphilis prevalence has been reported from 0.9 to 30.9% from 2000 to 2019. Specifically, the prevalence has been found 2.2%-30% in China [34-37], 1.1% -9% in Indonesia, 2.6%-4.46% in Bangladesh, 2.5% -14.1% in Myanmar, 5.5% in Combodia, 1.5%-4.8% in Nepal, 2.62%-17.5% in India, 1.65%-2.3% in Philippines, 0.9%-1.3% in Vietnam, 2.5% in Taiwan. A resurrection in Syphilis infection among MSM was also observed in the recent past, which has a strong association with HIV prevalence [38-41]. In many countries, it is observed that among MSM, 50% of the Syphilis cases are HIV positive [42, 43] which indicates a very high risk of re-infection among those MSM [44, 45]. Nonetheless, researches and well-defined reports on Syphilis as well as HIV-Syphilis co-infection are limited and data are inconsistent to estimate Syphilis infection rate among MSM in Asian countries [20].

Several countries with limited resources and evidence in Asia are facing a severe HIV epidemic. To control this epidemic, effective prevention programs are required [46]. Extended knowledge and evidence are essential for efficacious prevention programs. This study reveals the data gaps or paucity and provides new pieces of evidence. To our knowledge, this systematic review and meta-analysis is the first looking at the prevalence of HIV, Syphilis as well as HIV-Syphilis co-infection among MSM across Asia. Several systematic reviews and meta-analyses have been done, for example, in Asia [47] and central Asia [48] to observe the epidemic of HIV and sexual risk among men who have sex with men (MSM). Some meta-analyses have also been conducted in China to estimate HIV and Syphilis prevalence among MSM [49-52]. However, all of the studies conducted were country-specific, small-scale, and were backdated. Also, no study has been done yet to estimate the prevalence and inspect the trend of HIV, Syphilis, and HIV-Syphilis co-infection in Asia. For the first time, we are conducting a meta-analysis including all the countries of Asia (where a study was found) with the latest data to have a clear view of the prevalence and trend of HIV, Syphilis, and their co-infection among MSM in Asia.

## 2 Methods

The PRISMA statement and the MOOSE checklist [53-55] were followed in this systematic review and meta-analysis. The protocol of the review was not registared previously.

### 2.1 Search strategy and selection criteria

This is a comprehensive systematic review of published research papers on the prevalence of HIV, Syphilis, and co-infection of HIV and Syphilis among men who have sex with men (MSM). The search took place on January 5, 2021, by searching the following databases: PubMed, MEDLINE, and Google Scholar. The keywords “Prevalence”, “HIV”, “Syphilis”, “HIV and Syphilis co-infection”, “Men who have sex with men”, “MSM”, “Male sex workers”, “gay men” and all the countries in Asia (“India”, “Pakistan”, “Bangladesh”, “China”, and so on) and all possible combinations of them were used. For instance, one of the combinations is:

((((((((Prevalence [Title]) AND (HIV [Title])) OR (HIV and Syphilis co-infection [Title])) AND (Mem who have sex with men [Title])) OR (MSM [Title]) OR (China [Title]))))))))

EndNote (Version X8.1) reference management software was used to search. We mentioned the sources of articles or reports on MSM and HIV, Syphilis prevalence. We also searched websites of HIV surveillance and country-specific ministries of health. However, we considered the studies which were published between Jan 1, 2000, to December 30, 2020, and published only in English. All the identified research articles and reports were also transformed into EndNote software.

### 2.2 Inclusion/exclusion criteria

The studies were included if and only if the study population or part of the study population of the corresponding study is MSM or male sex workers or gay men or other men who had sex with men. The studies were excluded if the full text were not available, unrelated to the topic, studies without sufficient data such as did not report study design, study site as well as sample size or did not report at least one of the following information about MSM (1) prevalence of HIV, (2) Prevalence of Syphilis, and (3) prevalence of HIV-Syphilis co-infection. We also excluded Review Papers, Letters to the Editor, Brief Report, Editorial, Comment, Correspondence, Local reports, Ph.D. and Master thesis, Conference abstracts, and presentation.

### 2.3 Screening and extraction

Initially, the duplicates records and unrelated topics were removed by screening the titles and abstracts which was done by one author (SM). Two independent authors (AB, MI) randomly checked 10% of the initially screened records for accuracy and no inconsistencies were found. The screened records were considered for full-text review if titles and abstracts suggested that articles might have the relevant information. Then two reviewers (AI, SH) independently did the full-text review. The confusion or inconsistency was resolved by consulting with reviewer SH. Data extracted from eligible studies in a standard data extraction form (Excel file). Then data were also cross-checked for accuracy by the reviewer (SM) against the source.

### 2.4 Statistical analysis

All statistical analyses have been carried using statistical software STATA (version 16). The between study heterogeneity for the selected studies was assessed using the Q-test and I^2^ statistics with a 5% level of significant [55, 56]. In case of significant heterogeneity, we have applied a random effect model to estimate the pooled prevalence of HIV, syphilis, and their co-infection with 95% confidence intervals and the relative weight for each study. All the results of the meta-analysis were presented in either forest plots or tables. The funnel plot/ Egger’s test was used to identify the potential publication bias [53, 57]. The study location/ country, study duration, sampling method were considered for conducting subgroup analysis to observe the prevalence of them from different stratifications. The trends of prevalence of HIV, syphilis, and their co-infection have been inspected by assigning all selected studies into different groups based on the study duration.

## 3 Results

The Preferred Reporting Items for Systematic Reviews and Meta-Analyses (PRISMA) guidelines (Fig 1) were followed for collecting and reviewing the articles in this systematic review and meta-analysis. In the initial search, we identified a total of 2,704 articles including 168 surveillance reports. We removed 2,407 duplicates and title mismatches from the list. Then the screening was done for 465 records and we removed 194 studies. In the eligibility step, 205 studies were removed because at least one of the prevalence (HIV, Syphilis, Co-infection) was not reported, or study duration was not reported, or sampling method was not reported, study location was not reported, or study period was before 2000. Finally, 66 eligible studies were included in meta-analysis.

**Fig 1:**
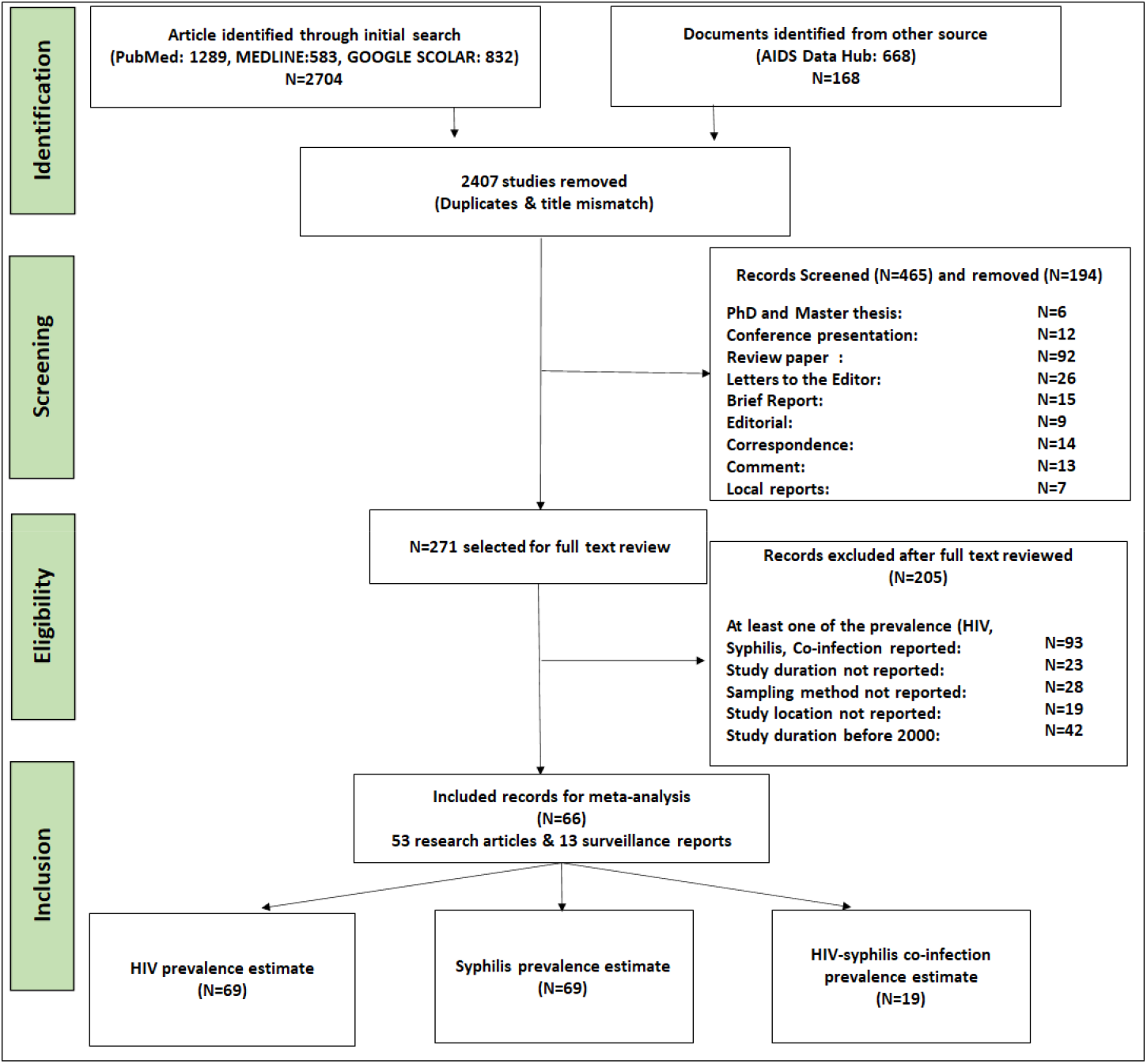
Flowchart describing the study selection approaches by following the PRISMA 2009 guidelines

The information that has been extracted from the finally selected studies is as follows: Name of the first author, year of publication, study location, study duration, study design, sampling methods, sample size or number of MSM tested for HIV, and the number of positive cases, number of MSM tested for Syphilis and number of positive cases, and the number of co-infected individuals (HIV-Syphilis).

### 3.1 Study characteristics

Table 1 describes the characteristics of the selected studies [4, 34, 35, 39, 40, 58-118]. Of the 40 studies that came from East Asia, 13 studies were from each region, South Asia, and South-East Asia. A total population of 128,510 for HIV and 129,090 for Syphilis has been covered under all eligible studies. Most of the eligible studies (52, including three consecutive sequential) are cross-sectional studies. Among the remaining studies, 13 are surveillance reports and one is a cohort study. A variety of sampling methods have been used among selected pieces of literature. The internet and venue-based sampling were used by 10.14% of selected studies, RDS (respondent-driven sampling) was considered in 27.54% of studies, Snowball sampling was considered in 15.94% of studies, Voluntary Counseling and Testing (VCT) was used by 5.80% of studies, Time location sampling was used by 5.80% of the studies, 11.59% of studies used Venue-day-time sampling (VDTS), whereas 5.80% studies used Convenience sampling methods, 10.14% of the studies used Multiple sampling methods and 7.25% of studies used other sampling methods (Community-based sampling, Long-chain referral sampling, Combination of RDS (respondent-driven sampling) and Snowball, Probability-based sampling, the combination of Cluster sampling and RDS. The average response rate is 92% (median= 99.3) with a minimum response rate is 43% and a maximum is 100%.

**Table 1:**
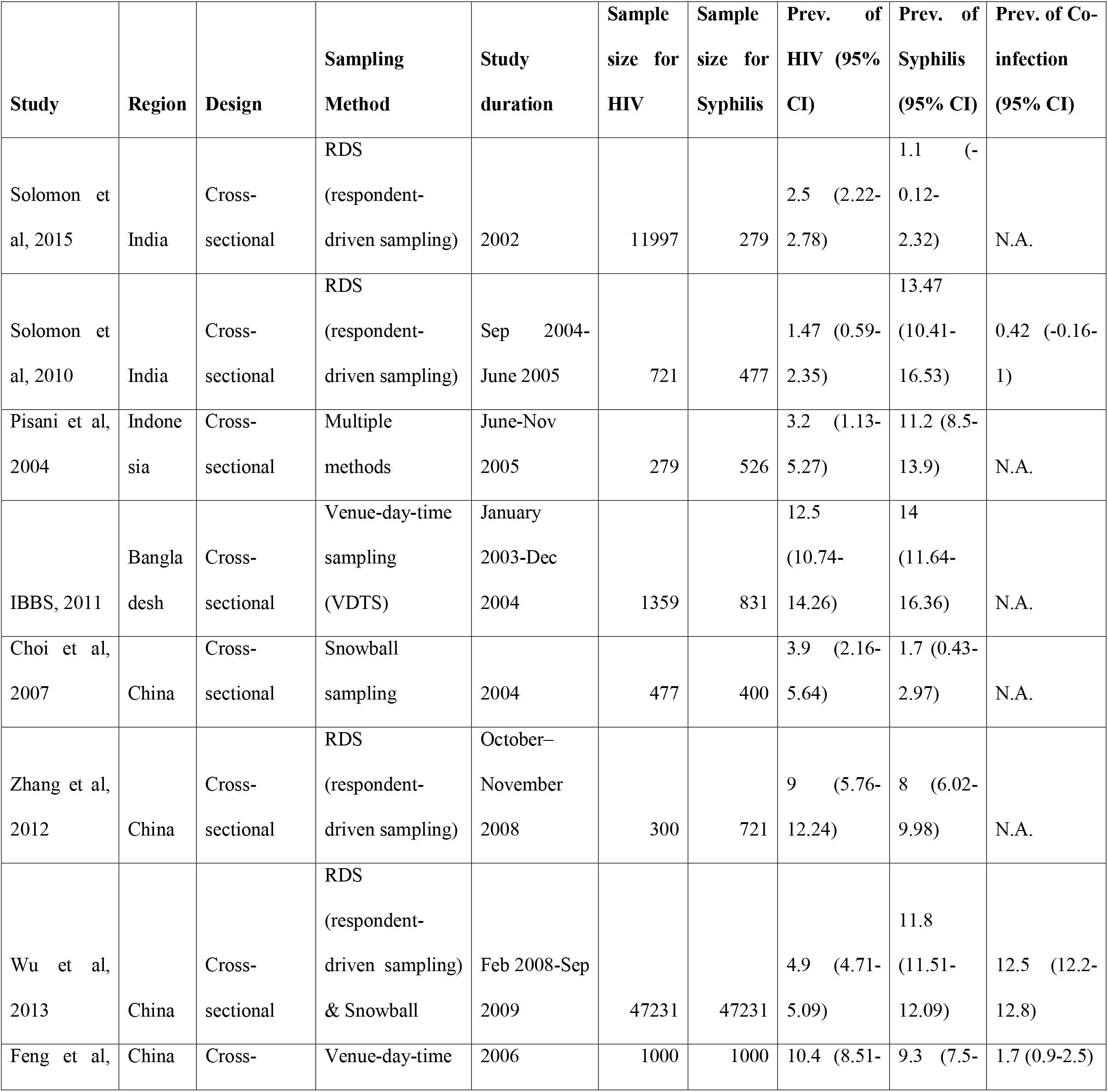
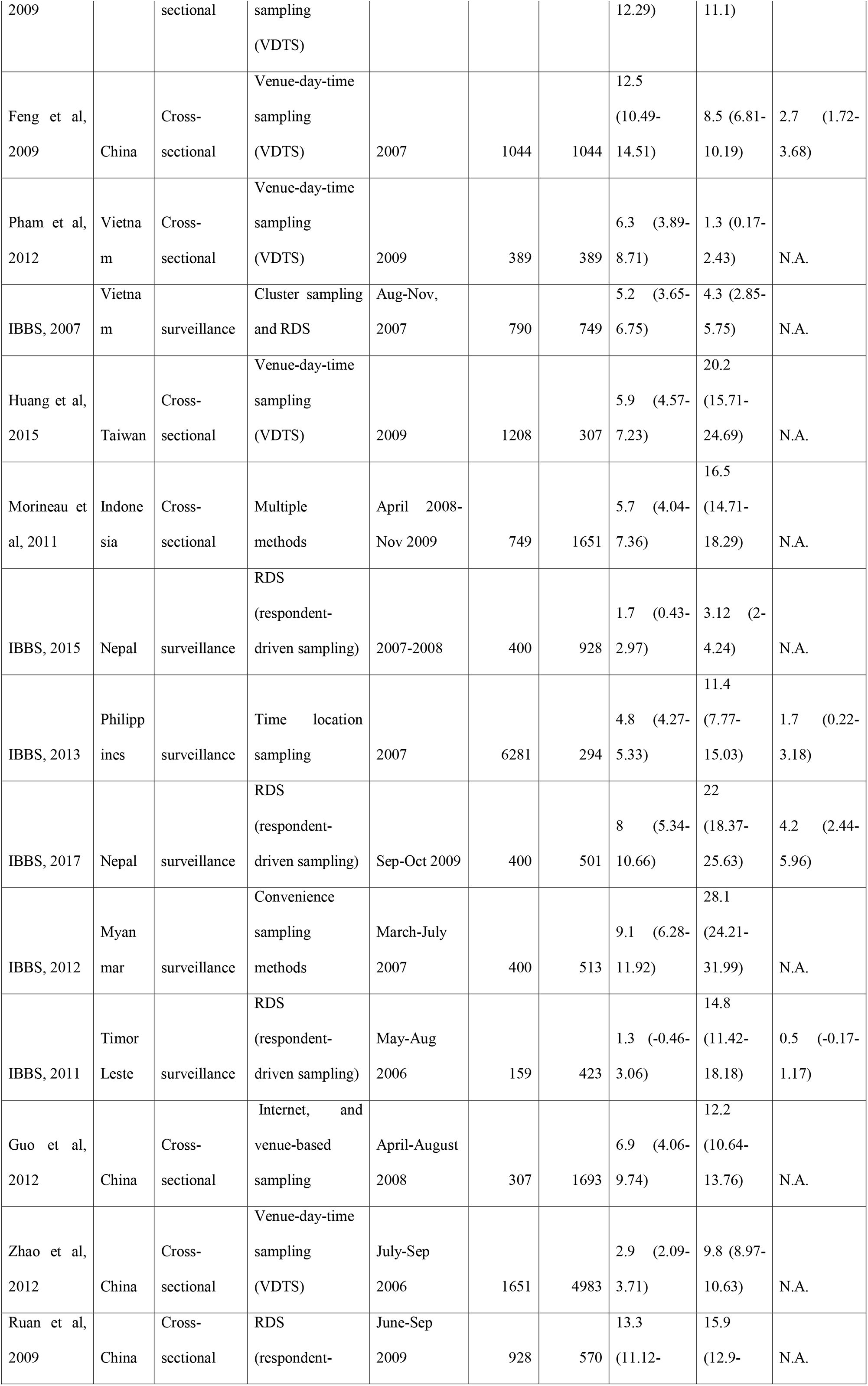
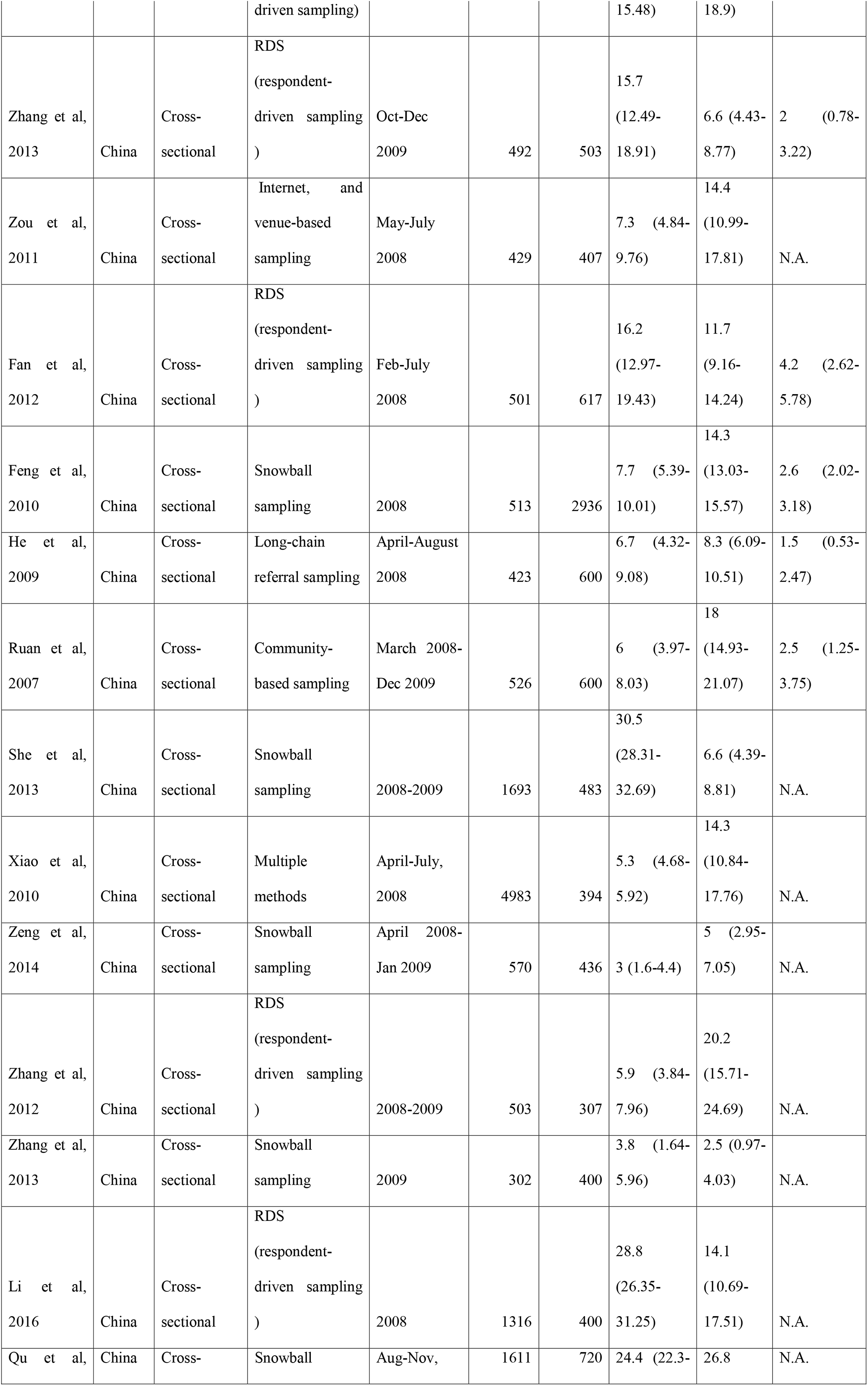
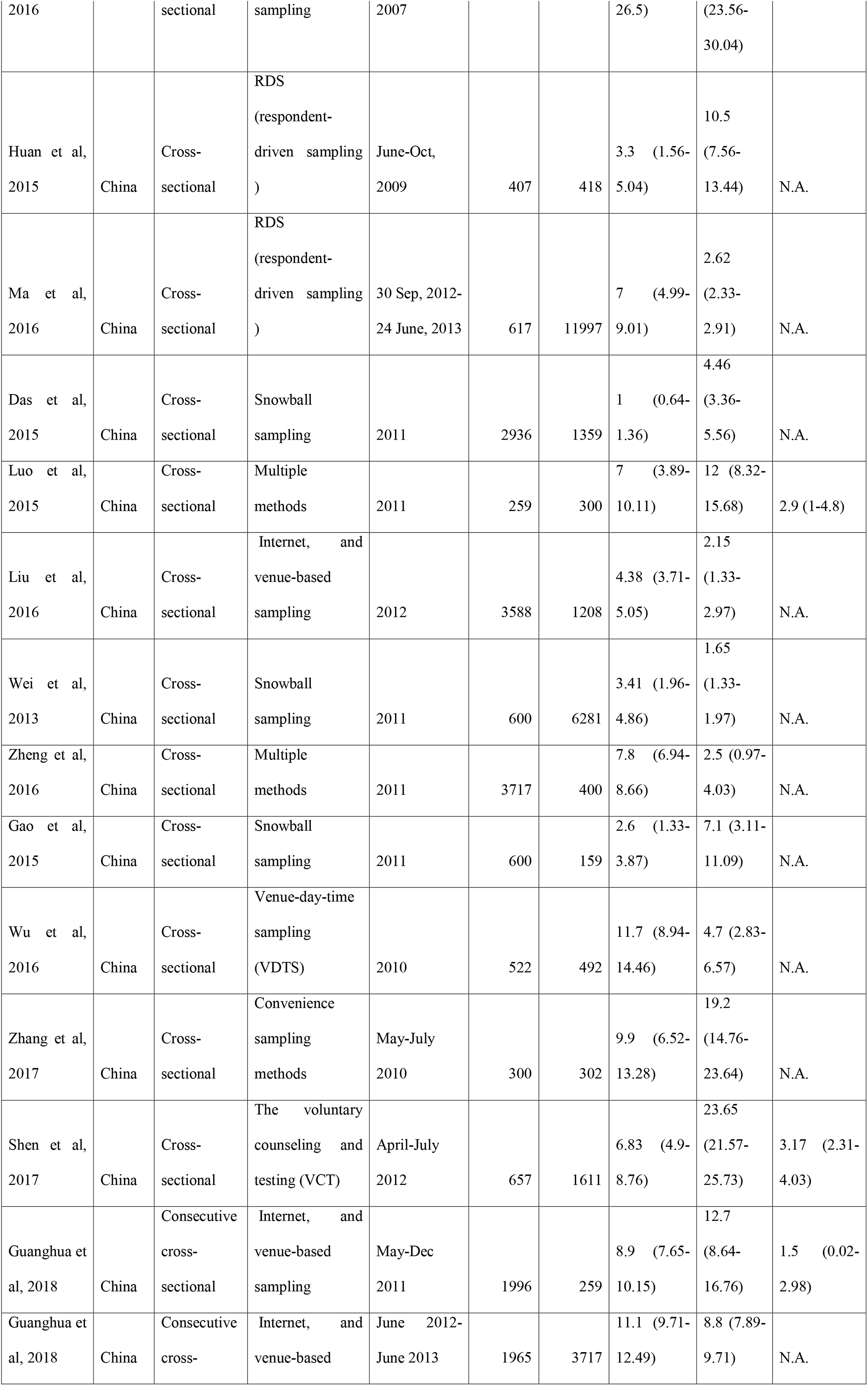
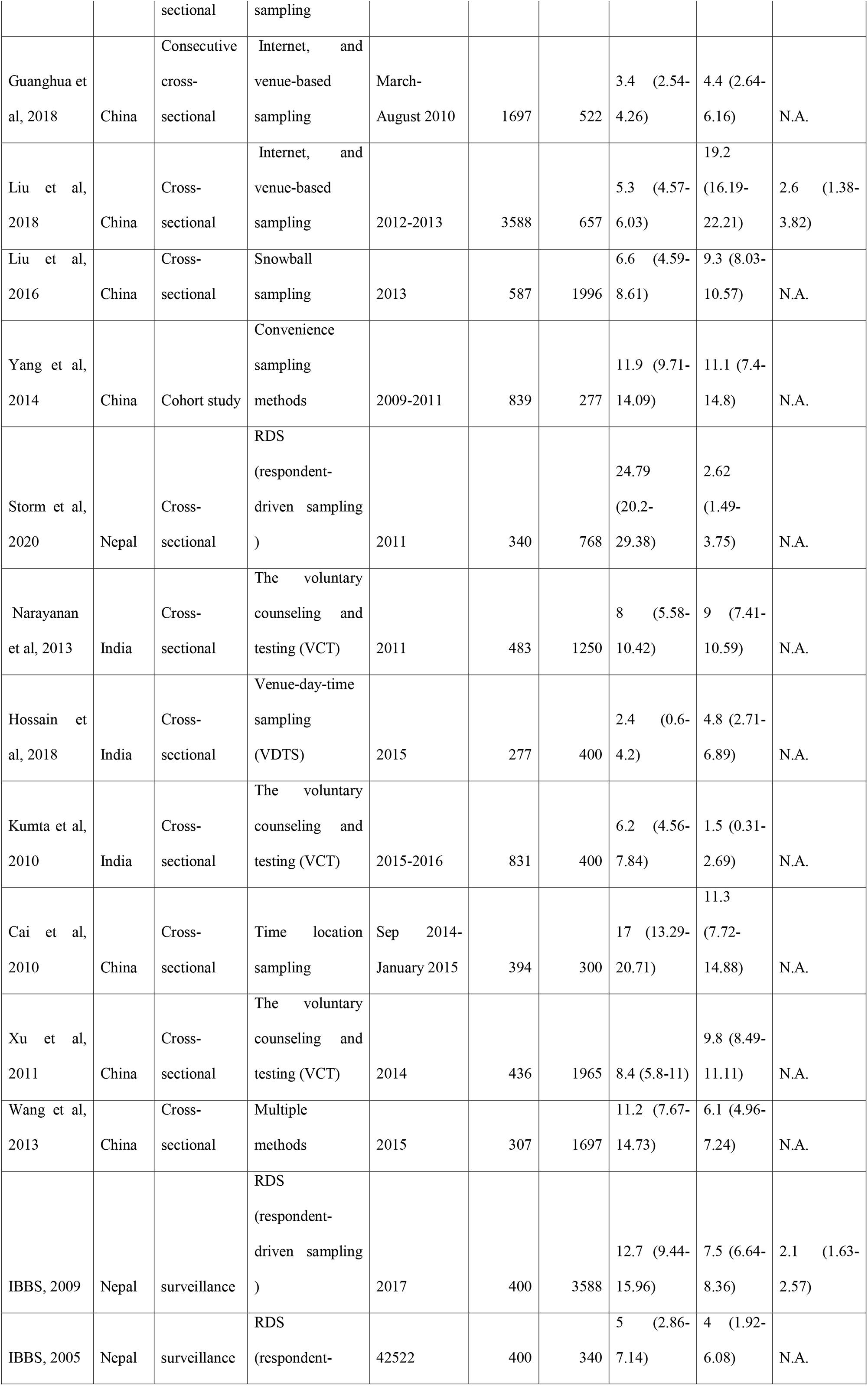
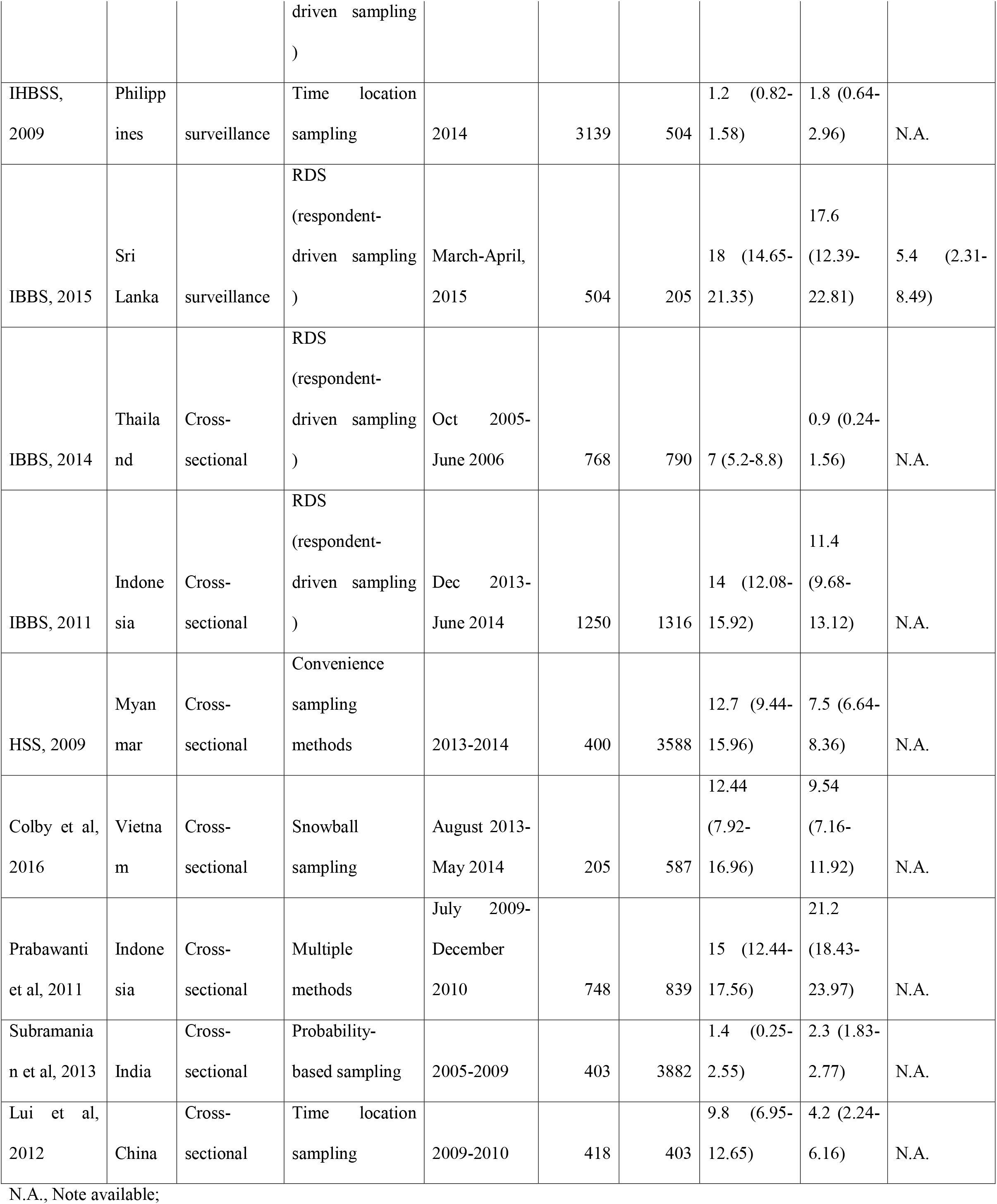
Features of the studies included in the Meta-analysis

### 3.2 Statistical heterogeneity and publication bias

The statistical tools *Q-*test and *I* ^2^ (%) with respective *p*-value were used to investigate the heterogeneity of the studies included in the analysis. We found statistically significant heterogeneity (*I*^2^ = 99.3, *Q* = *χ*^2^ (68)= 3414.66, *P* <0.01) for HIV (see Fig 2a) and Egger’s test showed that there was significant publication bias (z=5.7, *P*<0.001). Similarly, satistically significant heterogeneity (*I* ^2^ = 99.28%, *Q* = *χ*^2^ (68)= 5684.81, *P* <0.01) (see Fig 2b) and publication bias (z=7.7, *P*<0.001) were found for syphilis prevalence. Although, the heterogeneity (*I* ^2^ = 97.85%, *Q* = *χ*^2^ (17)= 3273.71, *P* <0.01) (see Fig 2c) for HIV and Syphilis co-infection were found statistically significant, Egger’s test showed that there was no significant publication bias (z=-0.04, *P*<0.001).

**Fig 2:**
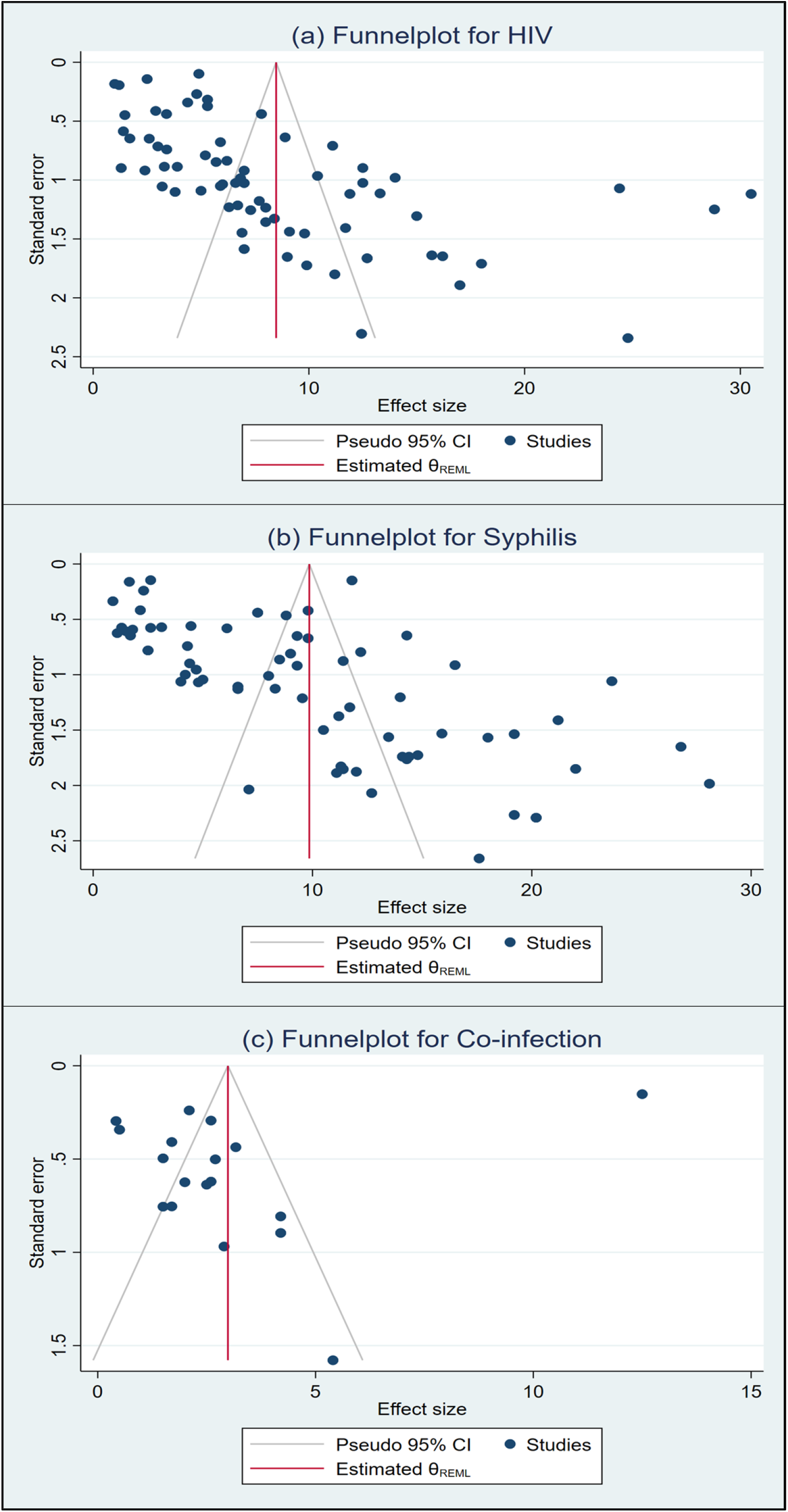
Funnel plot of result of the prevalence of HIV (a), Syphilis (b), and HIV and Syphilis co-infection (c) among MSM

### 3.3 Prevalence

The prevalence of HIV among MSM was assessed in 66 studies. The pooled prevalence of HIV was 8.48% (CI: 7.01-9.95, I^2^=99.35%) estimated from 69 estimates of 66 studies (presented in Fig 3). Similarly, 69 estimates in 66 studies were considered to estimate the overall prevalence of Syphilis among MSM in this meta-analysis. The estimated pooled prevalence of Syphilis was 9.86% (CI: 8.30-11.41, I^2^=99.28%) (shown in Fig 4). The prevalence of HIV and Syphilis co-infection was estimated in 17 studies out of 66 studies. The pooled prevalence of HIV-Syphilis co-infection was calculated as 2.99% (CI: 1.70 – 4.27, I^2^=97.85%) (Fig 5).

**Fig 3:**
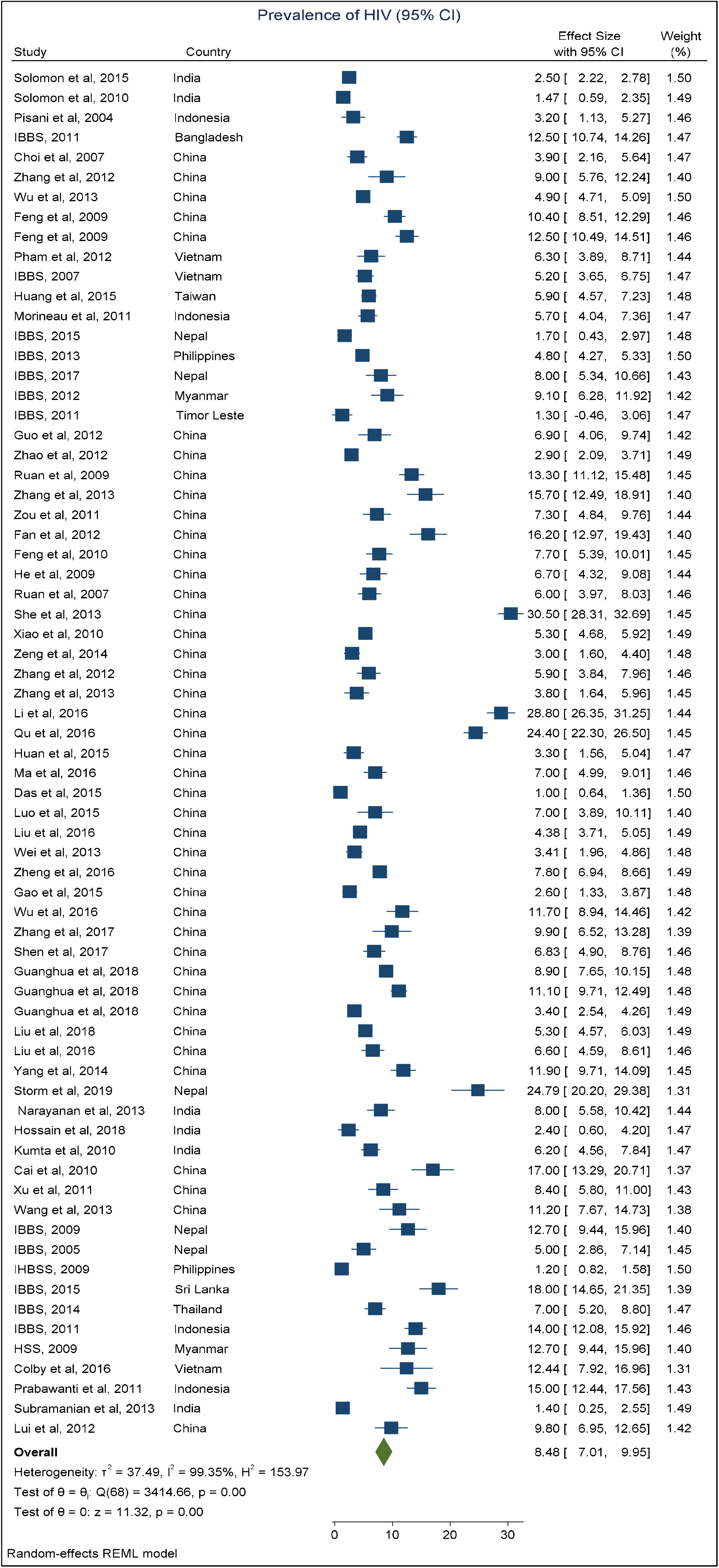
Forest plot showing the results of the pooled prevalence of HIV among MSM

**Fig 4:**
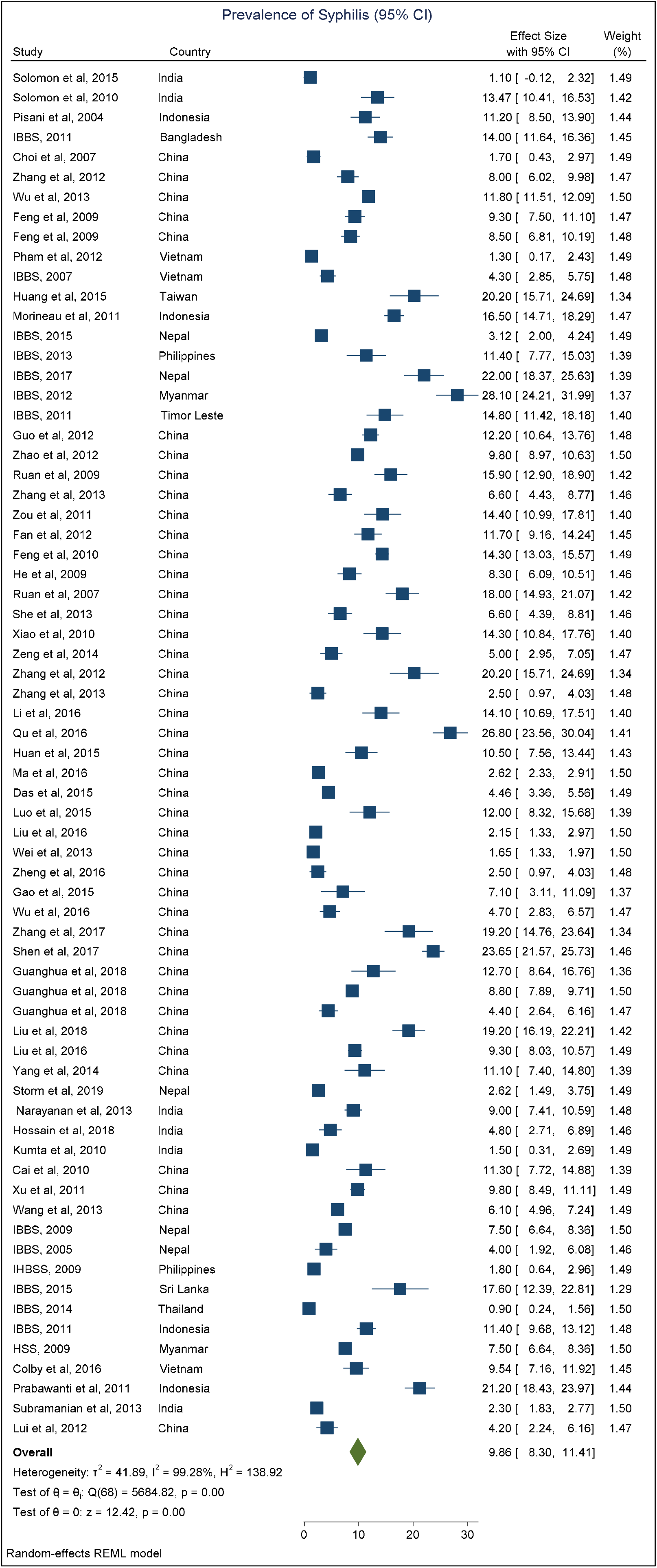
Forest plot showing the results of the pooled prevalence of Syphilis among MSM

**Fig 5:**
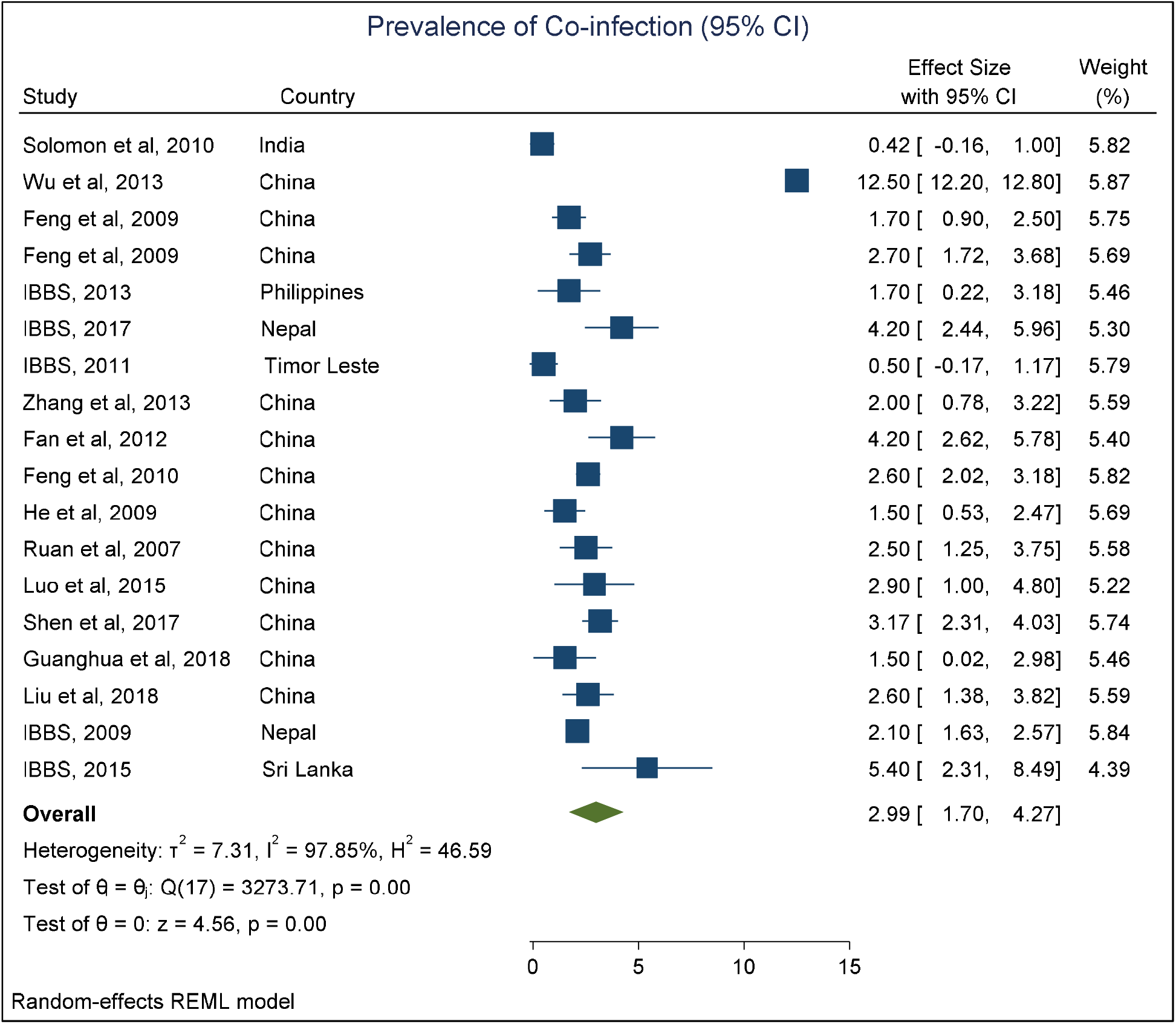
Forest plot showing the results of the pooled prevalence of HIV-Syphilis co-infection among MSM.

### 3.4 Trends of HIV, Syphilis, and HIV and Syphilis co-infection prevalence

The pooled estimate of HIV prevalence among MSM in Asia has an increasing trend with some fluctuations during 2002-2017. The prevalence of HIV was 2.50% (95% CI: 2.22-2.78) during 2002-2003 and then it gradually increased to 9.50% (95% CI: 6.25-12.74) during 2008-2009. Afterward, the prevalence fall to 7.83% (95% CI: 4.21-11.45) during 2010-2011 and stood at 6.82% (95% CI: 4.90-8.75) during 2012-2013. Then again the HIV prevalence increased and goes to its peak of 9.55% (95% CI: 3.84-15.25) during 2014-2015 and then reduced to 8.75% (95% CI: 1.20-16.29) during 2016-2017 (Fig 6). The overall HIV prevalence estimate irrespective of the subgroups is 8.31 (95% CI: 6.64-9.98) that is considered to be a significantly high level of between countries heterogeneity (1^2^ = 99.47%, p<0.001) (Fig 6).

**Fig 6:**
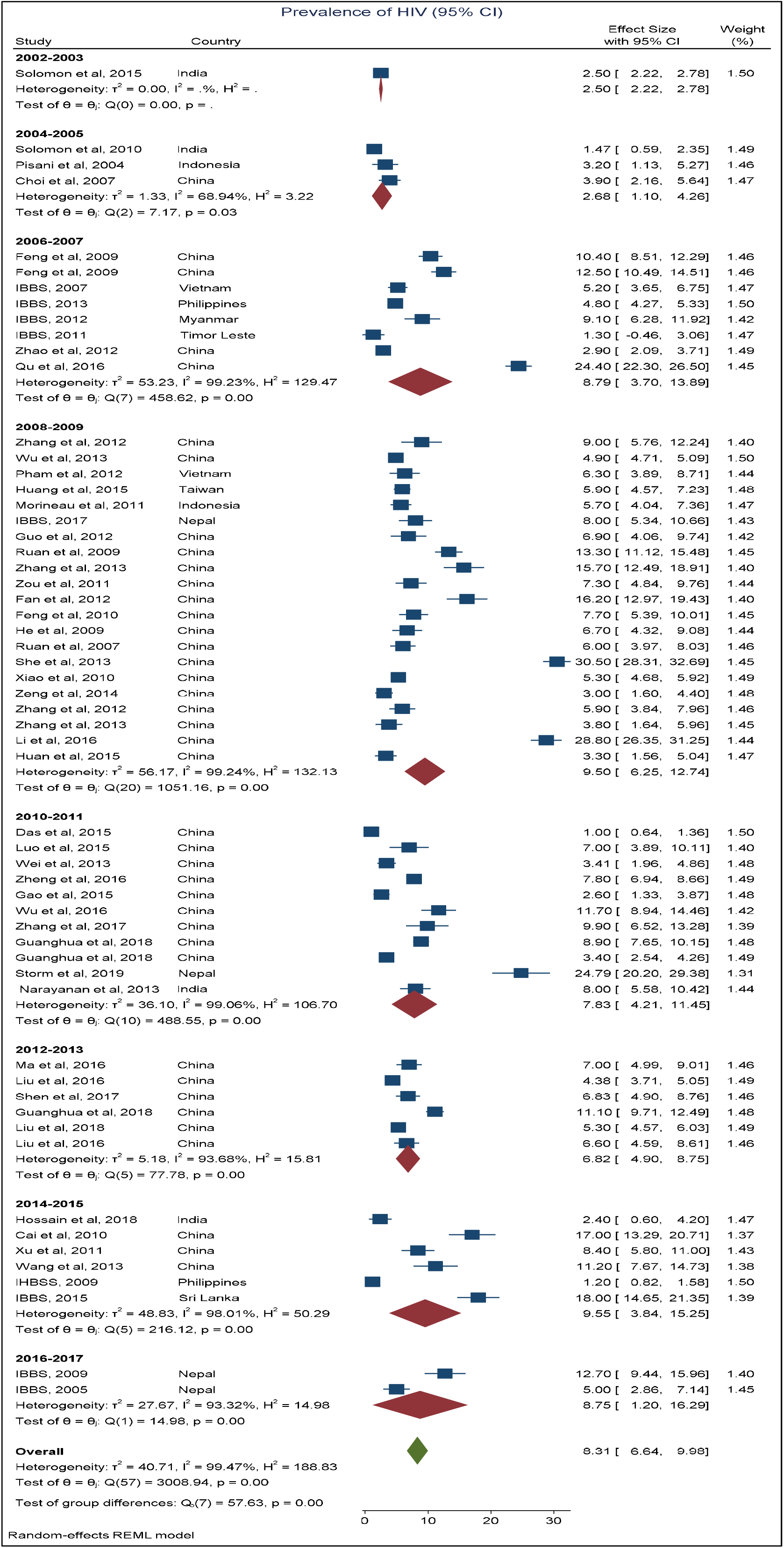
Forest plot showing changes in the prevalence of HIV over the time during 2002 to 2017

According to the findings from meta-analysis, the overall pooled estimate of Syphilis prevalence among MSM in Asia increased by study period during 2002-2007. The prevalence of Syphilis was 1.10% (95% CI: -0.12-2.32) in 2002-2003 period and increased to 8.68% (95% CI: 1.53-15.83) during 2004-2005 and reached its peak at 14.01% (95% CI: 8.01-20.02) during 2006-2007. After that, the prevalence of Syphilis kept going down and it was 5.88% (95% CI: 2.46-9.30) during 2016-2017. The overall Syphilis prevalence estimate regardless of the subgroup analysis is 10.25% (95% CI: 8.55-11.96) which indicates a significantly high level of between countries heterogeneity (1^2^ = 99.26%, p<0.001) (Fig 7).

**Fig 7:**
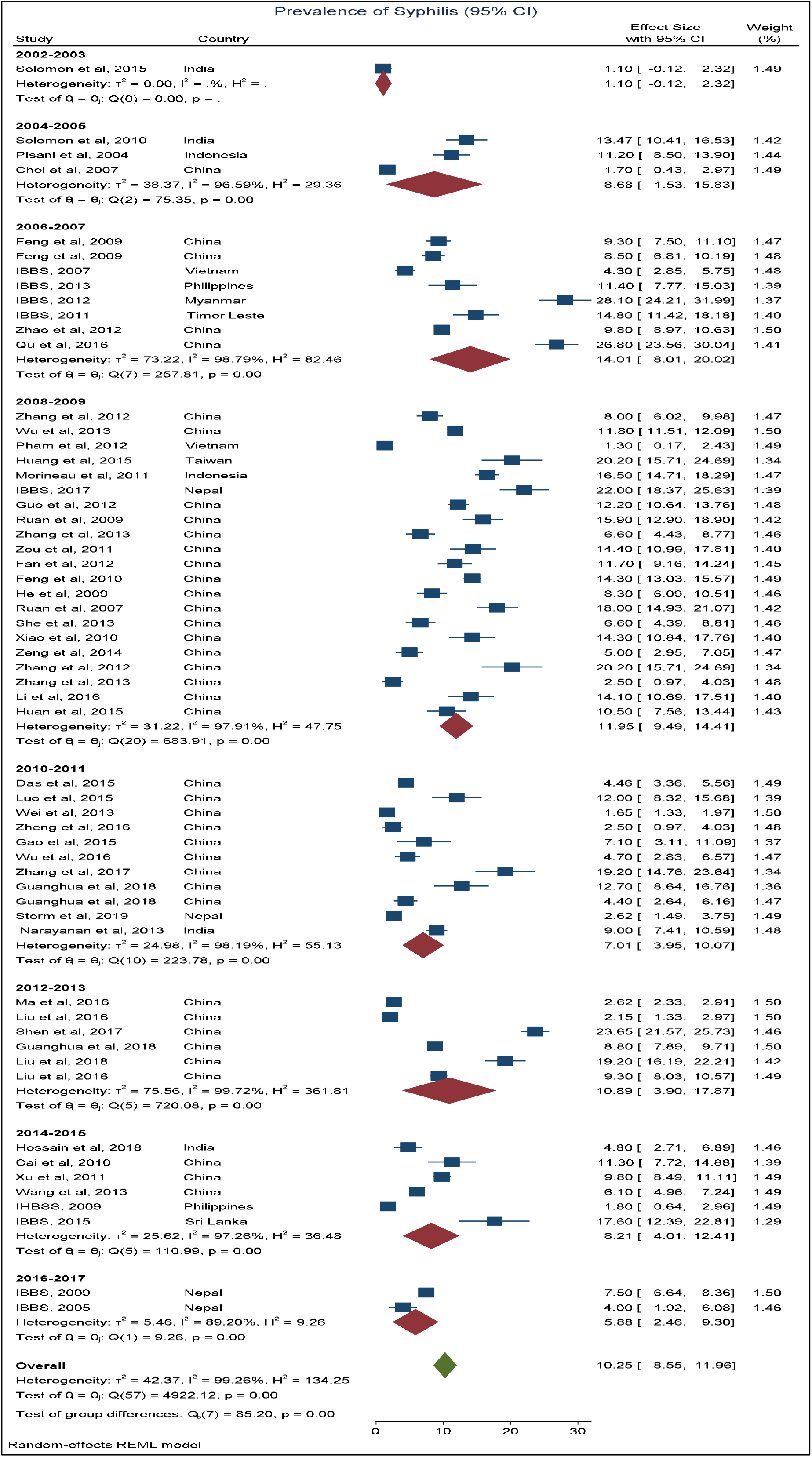
Forest plot showing changes in the prevalence of Syphilis over the time during 2002 to 2017

The findings from the meta-analysis show ups and downs among pooled estimates of HIV-Syphilis co-infection among MSM in Asia across all the studies over several periods. The pooled prevalence estimates of HIV-Syphilis co-infection was 0.42% (95% CI: -0.16-1.00) during the 2004-2005 terms. With some fluctuations, the prevalence estimate was found to be 2.10% (95% CI: 1.63-2.57) during 2016-2017. The overall HIV-Syphilis co-infection prevalence estimate irrespective of subgroup analysis is 2.99% (95% CI: 1.70-4.27) that indicating a considerably high level of between studies heterogeneity (1^2^ = 97.85%, p<0.001) (Fig 8).

**Fig 8:**
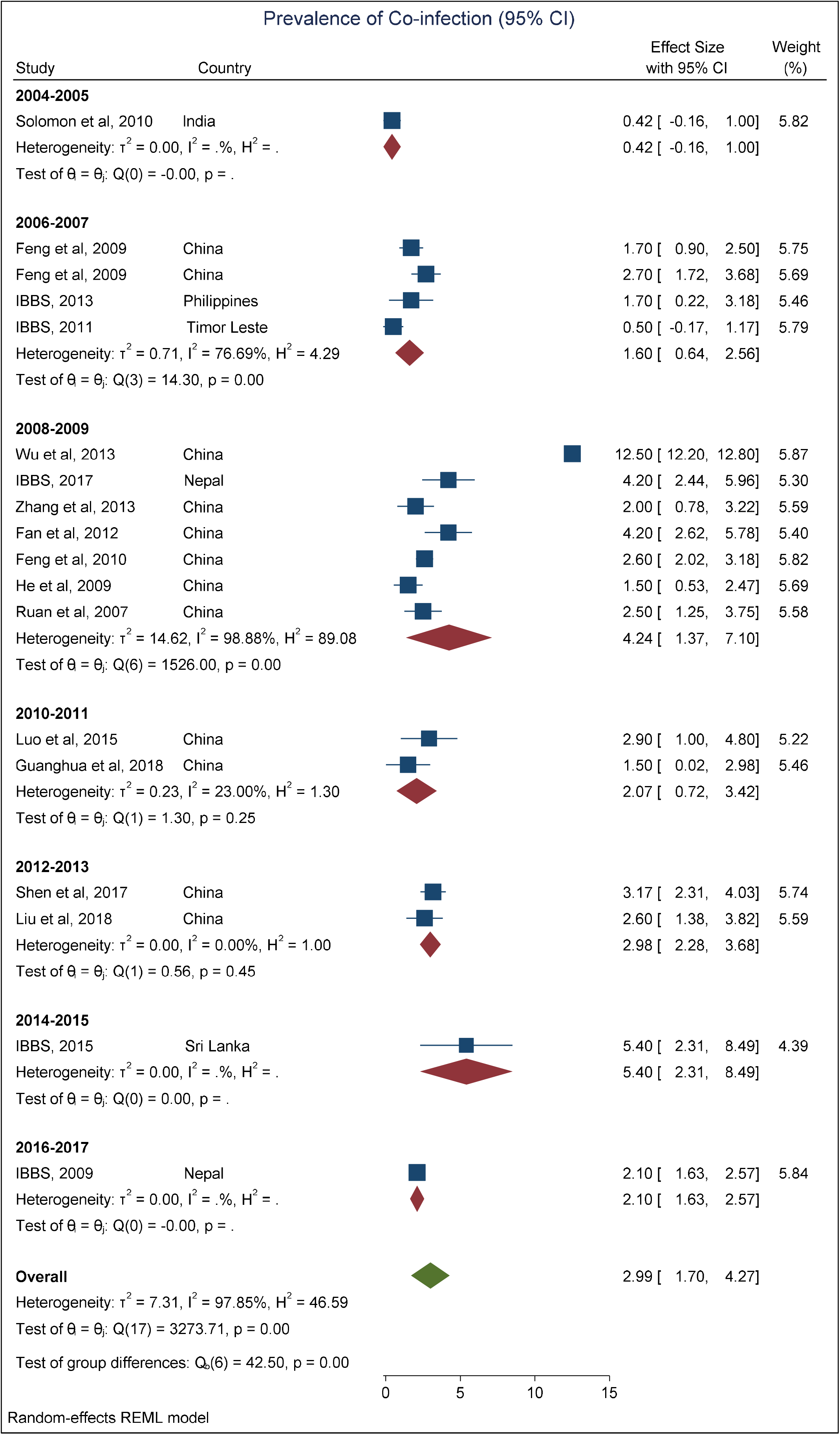
Forest plot showing changes in the prevalence of HIV and Syphilis co-infection over the time during 2002 to 2017

The HIV, Syphilis, and HIV-Syphilis co-infection prevalence estimates exhibited an upward trend consistent with study periods and HIV prevalence exhibited the most increasing trend where Syphilis prevalence exhibited the least increasing trend among the studies during 2002-2017 (Fig 9).

**Fig 9:**
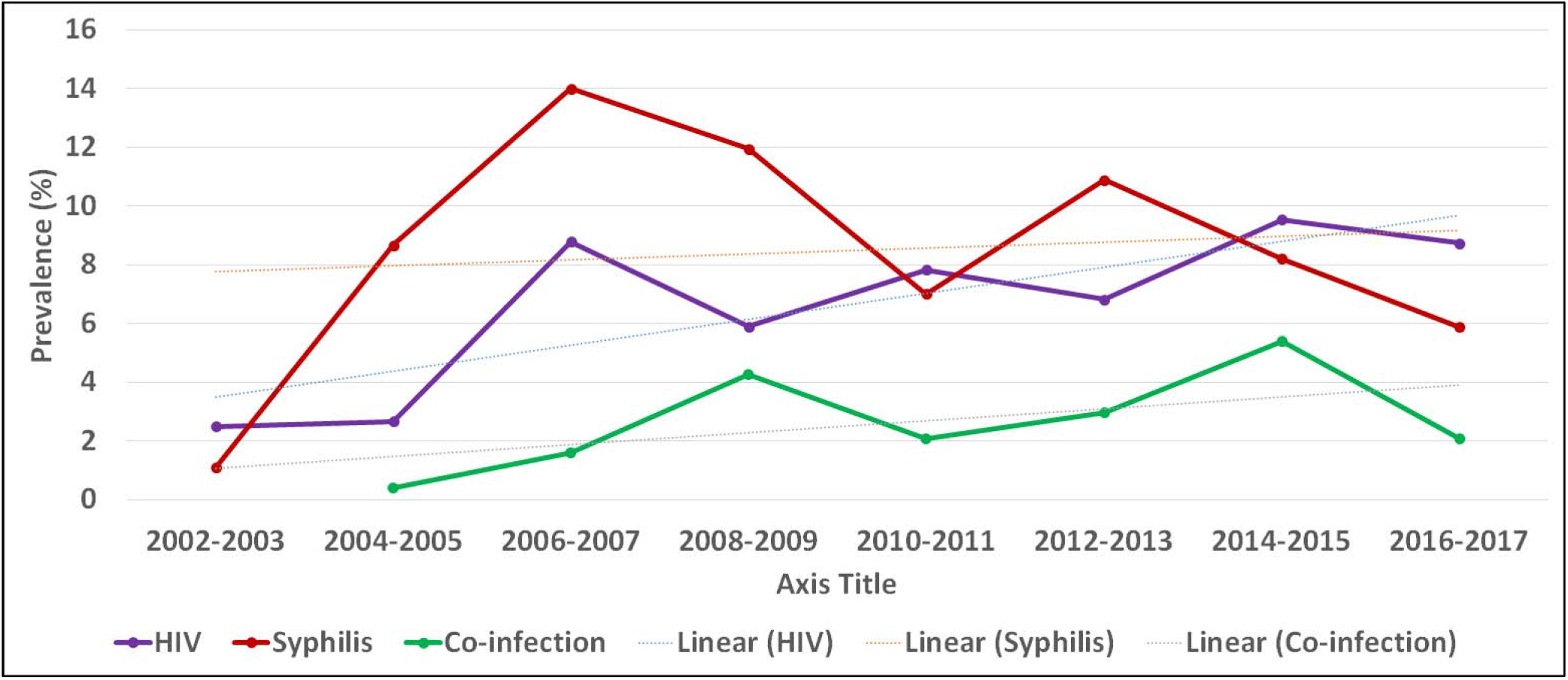
Trend in HIV, Syphilis, and HIV and Syphilis co-infection prevalence

### 3.5 Subgroup analysis based on Asian regions and countries

The prevalence of HIV, Syphilis, and HIV and Syphilis co-infection were investigated in the different territories of Asia. A total of 43 studies for HIV and Syphilis prevalence were included from the East Asian region and estimated overall HIV and Syphilis prevalence of 9% (95% CI: 7.06-10.93), presented in Fig 10 and 10.26% (95% CI: 8.43-10.10), presented in Fig 11 respectively. 13 studies were included in the analysis from the South Asian region and estimated overall HIV and Syphilis prevalence were 7.87% (95% CI: 4.08-11.66), presented in Fig 10 and 7.72% (95% CI: 4.12-11.32), presented in Fig 11 respectively. Totally 13 studies were included in this systematic review and meta-analysis from the Southeast Asian region and the estimated overall HIV prevalence was 7.38% (95% CI: 4.83-9.93), presented in Fig 10, and Syphilis prevalence was 10.65% (95% CI: 6.30-15.00), presented in Fig 11. A total of 12, 4, and 2 studies were included from the East Asian, South Asian, and Southeast Asian regions, and estimated overall HIV-Syphilis co-infection prevalence was 3.35% (95% CI: 1.60-5.09), 2.71% (95% CI: 0.64-4.78), and 0.91% (95% CI: 0.20-2.03) respectively presented in Fig 12. The highest prevalence of HIV in the Asian region was found at 30.50% (95% CI: 28.31-32.69) in the East Asian region and the lowest prevalence was 1% (95% CI: 0.64-1.36) also in the East Asian region. The highest prevalence of Syphilis was found at 28.10% (95% CI: 24.21-31.99) in the Southeast Asian region and the lowest prevalence was 0.90% (95% CI: 0.24-1.56) also in the Southeast Asian region. The highest prevalence of HIV-Syphilis co-infection was found at 12.50% (95% CI: 12.20-12.80) in the East Asian region and the lowest prevalence was 0.42% (95% CI: 0.16-1.00) in the South Asian region.

Table 2 displays the country-wise prevalence estimates of HIV, Syphilis, and HIV-Syphilis co-infection. From China, 42 studies were included in the analysis for the prevalence of HIV and Syphilis, 12 studies for HIV-Syphilis co-infection. Estimated overall prevalence of HIV, Syphilis, and HIV-Syphilis co-infection were 9.07% (95% CI: 7.10-11.05), 10.04% (95% CI: 8.22-9.08) and 3.55% (95% CI: 1.60-5.09) respectively in the Chinese studies. From India, 5 studies were included in the analysis for the prevalence of HIV and Syphilis, 1 study for HIV-Syphilis co-infection. Overall prevalence estimates of HIV, Syphilis, and HIV-Syphilis co-infection in the Indian studies were 3.52% (95% CI: 1.43-5.62), 5.23% (95% CI: 1.39-9.08) and 0.42% (95% CI: -0.16-1.00) respectively. From Indonesia, 4 studies were included and estimated overall HIV and Syphilis prevalence were 9.45% (95% CI: 3.67-15.22) and 15.04% (95% CI: 10.44-19.64) respectively. Four studies were included in the analysis for the prevalence of HIV and Syphilis, 2 studies for HIV-Syphilis co-infection from Nepal and estimated overall HIV, Syphilis, and HIV-Syphilis co-infection prevalence were 10.26% (95% CI: 2.53-17.99), 7.22% (95% CI: 0.77-14.67) and 2.97% (95% CI: 0.94-5.00) respectively. For a complete view of all countries, see Table 2.

**Table 2:** Subgroup analysis of HIV prevalence, Syphilis prevalence of HIV and Co-infection prevalence of HIV by country

| Region | HIV % (95% CI) (*i*<sup>2</sup>, Q(df.) P-value) | Syphilis % (95% CI) (*i*<sup>2</sup>, Q(df.) P-value) | Co-infection % (95% CI) (*i*<sup>2</sup>, Q(df.) P-value) |
| --- | --- | --- | --- |
| Bangladesh | 12.50 (10.74-14.26)* | 14.00 (11.64-16.36)* | -- |
| China | 9.07 (7.10-11.05)<br>(*i*<sup>2</sup> = 99.34, Q(41) = 2312.93, p-value < 0.001) | 10.04 (8.22-9.08)<br>(*i*<sup>2</sup> = 99.23, Q(41) = 4221.71, p-value < 0.001) | 3.55 (1.60-5.09)<br>(*i*<sup>2</sup> = 98.08, Q(11) = 2199.10, p-value < 0.001) |
| India | 3.52 (1.43-5.62)<br>(*i*<sup>2</sup> = 96.11, Q(5) = 48.21, p-value < | 5.23 (1.39-9.08)<br>(*i*<sup>2</sup> = 98.29, Q(5) = 125.03, p-value < | 0.42 (-0.16-1.00)* |
|  | 0.001) | 0.001) |  |
| Indonesia | 9.45 (3.67-15.22)<br>($I^2 = 96.97$, Q(3) = 92.22, p-value < 0.001) | 15.04 (10.44-19.64)<br>($I^2 = 94.51$, Q(3) = 45.62, p-value < 0.001) | -- |
| Myanmar | 10.80 (7.28-14.33)<br>($I^2 = 62.67$, Q(1) = 2.68, p-value = 0.10) | 17.7 (-2.48-37.90)<br>($I^2 = 99.03$, Q(1) = 102.70, p-value < 0.001) | -- |
| Nepal | 10.26 (2.53-17.99)<br>($I^2 = 98.09$, Q(4) = 123.71, p-value < 0.001) | 7.22 (0.77-14.67)<br>($I^2 = 99.26$, Q(4) = 144.15, p-value < 0.001) | 2.97 (0.94-5.00)<br>($I^2 = 80.49$, Q(1) = 5.13, p-value = 0.02) |
| Philippines | 3.00 (-0.53-6.52)<br>($I^2 = 99.15$, Q(1) = 117.16, p-value < 0.001) | 6.44 (-2.96-15.84)<br>($I^2 = 95.89$, Q(1) = 24.34, p-value < 0.001) | 1.70 (0.22-3.18)* |
| Sri Lanka | 18.00 (14.65-21.35)* | 17.60 (12.39-22.81)* | 5.40 (2.31-8.49)* |
| Taiwan | 5.90 (4.57-7.23)* | 20.20 (15.71-24.69)* | -- |
| Thailand | 7.00 (5.20-8.80)* | 0.90 (0.24-1.56)* | -- |
| Timor-Leste | 1.30 (-0.46-3.06)* | 14.80 (11.42-18.18)* | 0.50 (-0.17-1.17)* |
| Vietnam | 7.51 (3.56-11.45)<br>($I^2 = 85.76$, Q(2) = 8.88, p-value = 0.01) | 4.95 (0.29-9.61)<br>($I^2 = 96.27$, Q(2) = 40.43, p-value < 0.001) | -- |

\*refers only one studies were included in this subgroup; - refers there is no studies were considered in this subgroup; CI, Confidence Interval

**Fig 2:**
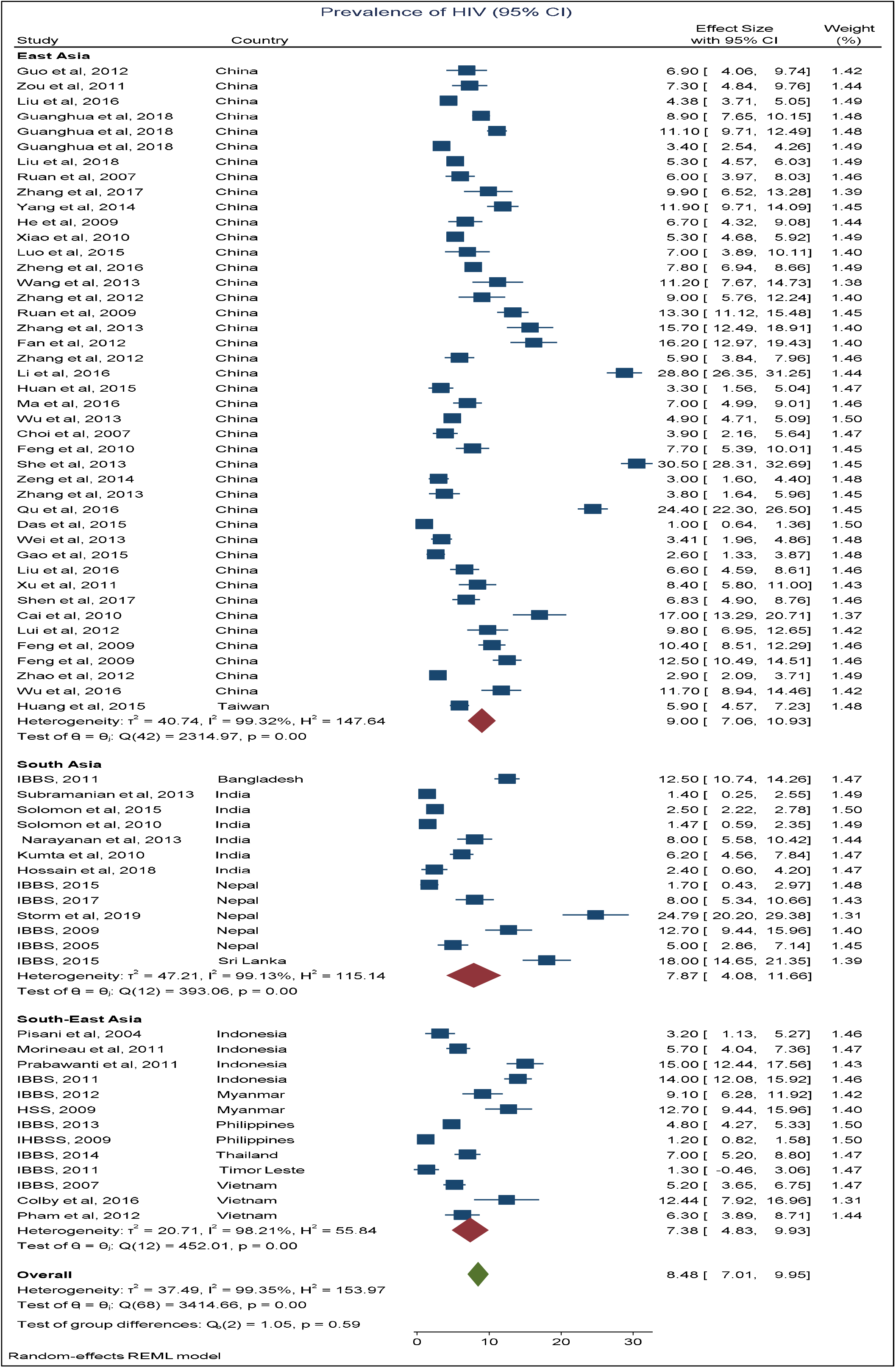
Forest plot showing regional disparities in the prevalence of HIV from 2002 to 2017 in Asia

**Fig 3:**
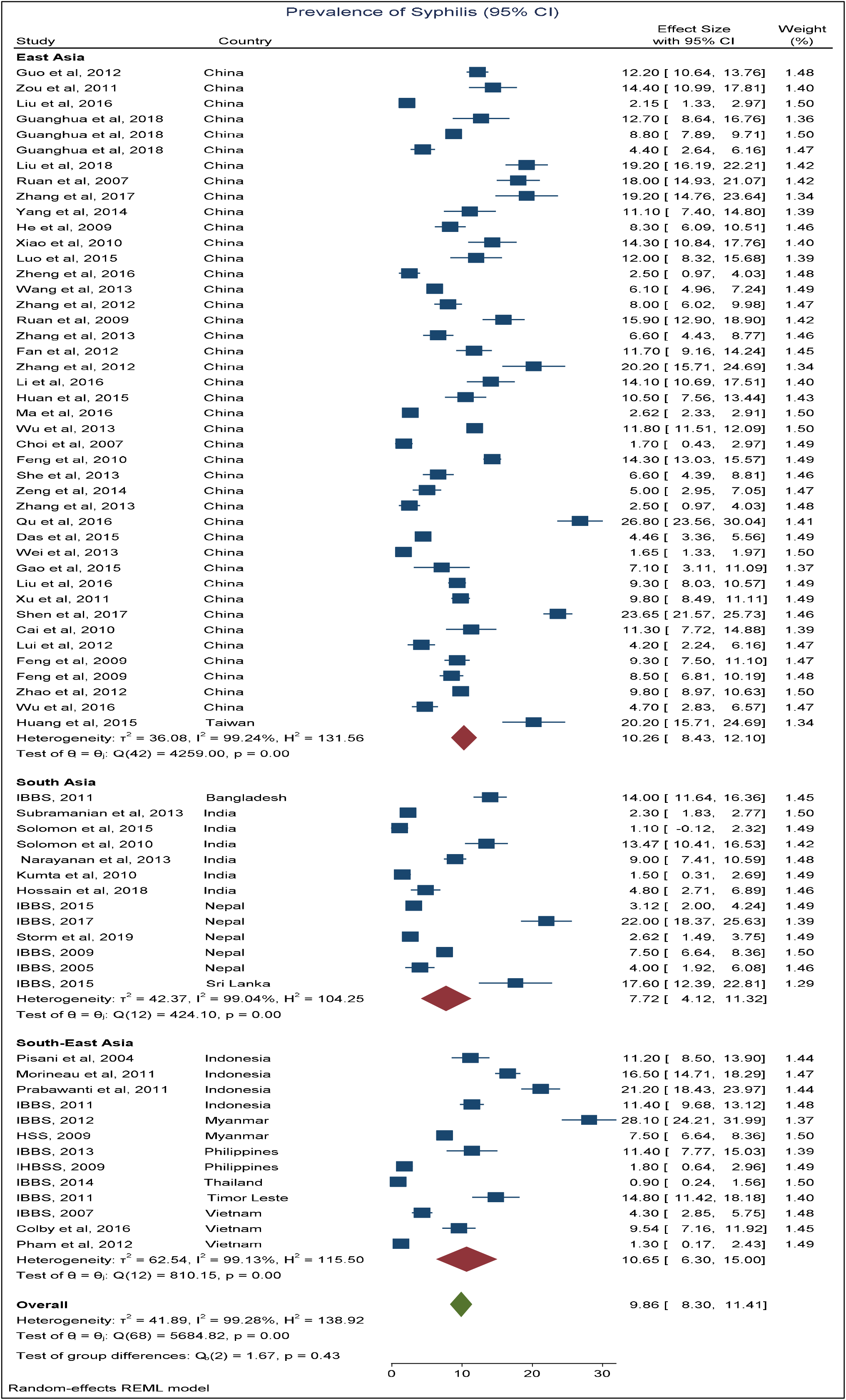
Forest plot showing regional disparities in Syphilis of HIV during 2002 to 2017 in Asia

**Fig 4:**
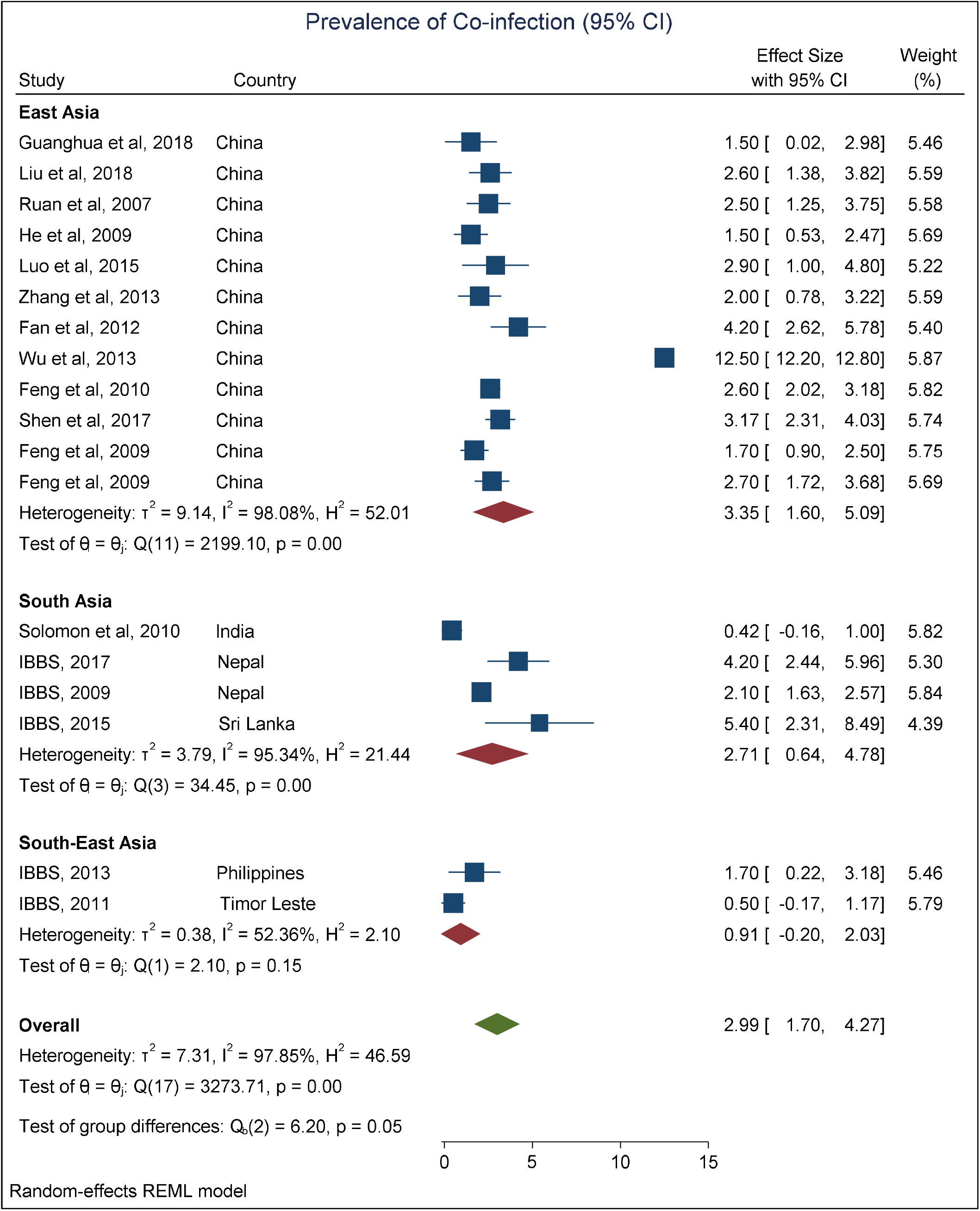
Forest plot showing regional disparities in the prevalence of HIV and Syphilis co-infection from 2002 to 2017 in Asia

### 3.6 Subgroup analysis based on sampling methods

Table 3 exhibits the prevalence estimates of HIV, Syphilis, and HIV-Syphilis co-infection by different sampling methods used. Internet and venue-based sampling method was used to estimate the prevalence of HIV and Syphilis in 7 studies, for HIV-Syphilis co-infection in 2 studies among MSM. Here, estimated overall HIV, Syphilis, and HIV-Syphilis co-infection prevalence were 5.25% (95% CI: 2.34-8.17), 8.67% (95% CI: 4.65-12.69) and 6.46% (95% CI: -5.38-18.30) respectively. The estimated overall HIV, Syphilis, and HIV-Syphilis co-infection prevalence based on community-based sampling method were 7.08% (95% CI: 4.46-9.71), 10.00% (95% CI: 3.07-16.94) and 2.70% (95% CI: 1.72-3.68) respectively. See Table 3 for other methods.

**Table 3:** Subgroup analysis of HIV prevalence, Syphilis prevalence of HIV, and Co-infection prevalence of HIV by sampling method

| Sampling Methods | HIV % (95% CI) ($I^2$, Q(df.) P-value) | Syphilis % (95% CI) ($I^2$, Q(df.) P-value) | Co-infection % (95% CI) ($I^2$, Q(df.) P-value) |
| --- | --- | --- | --- |
| Internet and venue-based sampling | 5.25 (2.34-8.17) | 8.67 (4.65-12.69) | 6.46 (-5.38-18.30) |
| | ($I^2 = 99.49$, $Q(6) = 323.65$,<br>p-value < 0.001) | ($I^2 = 98.49$, $Q(6) = 502.63$,<br>p-value < 0.001) | ($I^2 = 99.92$, $Q(1) = 1316.53$,<br>p-value < 0.001) |
| Community-based sampling | 7.08 (4.46-9.71)<br>($I^2 = 91.38$, $Q(4) = 38.53$, p-value < 0.001) | 10.00 (3.07-16.94)<br>($I^2 = 98.82$, $Q(4) = 250.91$,<br>p-value < 0.001) | 2.70 (1.72-3.68)* |
| Long-chain referral sampling | 1.70 (0.43-2.97)* | 3.12 (2.00-4.24)* | -- |
| Multiple methods | 6.51 (3.49-9.54)<br>($I^2 = 97.20$, $Q(6) = 110.59$,<br>p-value < 0.001) | 16.18 (11.39-20.96)<br>($I^2 = 96.97$, $Q(6) = 130.78$,<br>p-value < 0.001) | 2.02 (-0.09-4.12)<br>($I^2 = 87.85$, $Q(2) = 15.66$, p-value < 0.001) |
| RDS (respondent-driven sampling) | 9.17 (5.15-13.18)<br>($I^2 = 99.57$, $Q(18) = 1773.44$, p-value < 0.001) | 9.59 (6.45-12.72)<br>($I^2 = 99.46$, $Q(18) = 965.66$,<br>p-value < 0.001) | 2.59 (1.81-3.38)<br>($I^2 = 58.34$, $Q(4) = 8.78$, p-value = 0.07) |
| Snowball sampling | 9.62 (6.55-12.69)<br>($I^2 = 97.74$, $Q(10) = 219.38$,<br>p-value < 0.001) | 11.22 (7.19-15.25)<br>($I^2 = 98.34$, $Q(10) = 424.59$,<br>p-value < 0.001) | 2.58 (1.67-3.49)<br>($I^2 = 45.75$, $Q(2) = 3.71$, p-value = 0.16) |
| Voluntary counseling and testing (VCT) | 8.34 (2.42-14.27)<br>($I^2 = 96.46$, $Q(3) = 52.39$, p-value < 0.001) | 6.71 (2.29-11.14)<br>($I^2 = 96.18$, $Q(3) = 94.91$, p-value < 0.001) | -- |
| Time location sampling | 7.33 (2.00-12.65)<br>($I^2 = 96.42$, $Q(3) = 86.86$, p-value < 0.001) | 4.89 (2.39-7.40)<br>($I^2 = 94.25$, $Q(3) = 62.77$, p-value < 0.001) | 2.10 (1.63-2.57)* |
| Venue-day time sampling (VDTS) | 11.17 (7.46-14.87)<br>($I^2 = 99.03$, $Q(1) = 102.70$,<br>p-value < 0.001) | 9.15 (4.21-14.09)<br>($I^2 = 99.03$, $Q(1) = 102.70$,<br>p-value < 0.001) | 5.40 (2.31-8.49)* |
| Other | 11.03 (6.36-15.70)<br>($I^2 = 95.35$, $Q(7) = 240.12$,<br>p-value < 0.001) | 9.92 (5.62-14.22)<br>($I^2 = 99.39$, $Q(7) = 459.31$,<br>p-value < 0.001) | 1.79 (1.12-2.46)<br>($I^2 = 0.00$, $Q(1) = 0.16$, p-value = 0.69) |

\*refers only one study was included in this subgroup; - refers no studies were considered in this subgroup; CI, Confidence Interval

## 4 Discussion

To our knowledge, this meta-analysis provides the first comprehensive quantitative analysis of the prevalence of HIV, Syphilis, and their co-infection and also inspected the trends (2002-2017, with a 5-year interval) of HIV, Syphilis, and their co-infection among MSM across Asia. This review paper also illustrates the heterogeneity of the epidemiology of HIV, and Syphilis and their co-infection over different geographical regions and countries.

The Burden of HIV, Syphilis infection and co-infection of HIV and Syphilis is disproportionately high among high-risk populations especially among men who have sex with men (MSM). Globally, this is inspected that the Syphilis epidemic has a strong association with the increase of the HIV incidence among MSM [38, 40, 41]. Despite the early victories at abridging the spread of HIV in some regions like Thailand and Cambodia, the HIV/AIDS epidemic proceeds to develop in the pockets across Asia. The studies in the near past demonstrated that the incidence of sexually transmitted infections (STI) rapidly increases among risk groups and the general population. The scenario of the HIV/AIDS epidemic varies from nation to nation, with countries in East Asia carrying a heavy burden of the illness. HIV/AIDS also differs considerably by the political, cultural, and economical diversity of the nations in Asia [46].

A total of 66 studies including 13 surveillance reports were found eligible from different countries. A total of 69 prevalence estimates from those 66 studies provide the pooled prevalence of HIV of 8.48% with a combined total of 128,510 participants, Syphilis of 9.86% with a combined total of 129,090 respondents. In addition, 17 studies with a combined total of 61,802 participants showed the prevalence of HIV-Syphilis co-infection at 2.99% across Asia. A higher prevalence of HIV was observed in 2018 in Eastern and Southern Africa (13.2%), western and central Africa (13.7%), and Latin America (12.6%). Comparatively lower prevalence was found in 2018 in Eastern Europe and Central Asia (6.2%), Western and Central Europe and North America (6.7%), Asia and the Pacific (4.9%) [119]. A global systematic review estimated a higher prevalence of Syphilis in Latin America and the Caribbean (10.6%) and lower in Australia and New Zealand (1.9%) during 2000-20 compared to our estimates[120].

Despite a relatively lower prevalence of HIV, Syphilis, and their co-infection in 2002-2003, the overall pooled estimate of HIV prevalence among MSM in Asia substantially fluctuated by the study period during 2000-2017. The prevalence of HIV, Syphilis and their co-infection among MSM had an upward trend across Asia (Fig 8 and 9). This finding is consistent with previous researches [49, 51, 121]. The highest prevalence of HIV (9.55%), and co-infection (5.40%) was found during 2014-15, and the lowest prevalence estimates of HIV (2.50%) and co-infection (0.42%) were observed during 2002-03, and 2004-2005, respectively. However, the highest prevalence of Syphilis was at 14.01% during 2006-07 and the lowest prevalence was 1.10% during 2002-03. The spread might be linked to MSM’s unique position in the world. The mainstream public still has a hard time accepting homosexuality. As a result, MSM and women marry often, and MSM may function as a conduit for HIV transmission to other MSM and the general population.

The stratified analyses among geographical regions (East Asia, South Asia, and South-East Asia) showed that the prevalence of HIV, Syphilis, and their co-infection varied by region. In East Asia, the overall prevalence of HIV and Syphilis were 9% and 10.26%, respectively. China exhibits the highest and lowest prevalence in all three cases. A study was conducted in China to find the frequency of bisexual activity among MSM and was found to be as high as 31.2 percent [122] which is consistent with our findings. Another systematic review in China observed an increased risk of HIV acquisition after Syphilis exposure [123]. In South Asia, the overall prevalence estimates of HIV and Syphilis were 7.87%, and 7.72%, respectively. Among all South Asian countries, the highest prevalence of HIV and Syphilis was found in Nepal, on the other hand, the lowest was in India. Besides, the highest prevalence of co-infection was found in Sri Lanka. Although HIV and Syphilis cases are declining as compared to previous decades, according to UNAIDS, Nepal has still 30,000 HIV-positive adults estimated in 2020 [124]. High-risk groups in Nepal include MSM which is consistent with our study [125]. Sex worker trafficking across borders is a serious issue in Nepal, leading to rising HIV rates. With a greater focus on a two-way flow paradigm, the old idea of HIV, and Syphilis entering Nepal from India is being questioned [126]. Although Sri Lanka is thought to have an extremely low HIV prevalence rate [125], in our study, we found a higher risk of co-infection among MSM in Sri Lanka. The prevalence of Syphilis and their co-infection among MSM was 5% in Sri-Lanka estimated in 2020 [127] which is consistent with our findings of overall co-infection of 5.40% in Sri Lanka.

In Southeast Asia, the pooled prevalence of HIV and Syphilis are 7.38%, and 10.65%, respectively. The highest (and lowest) prevalence of HIV and Syphilis in this region are in Indonesia (Philippines), and in Myanmar (Thailand), respectively which is consistent with previous studies where MSM shown to have a high and growing HIV prevalence rate, particularly in India, Indonesia, Myanmar, and Thailand [127]. In these regions, a sizable proportion of the MSM population engages in high-risk HIV behaviors. Multiple male sexual partners of all sorts – regular, casual, and paid – combined with irregular condom usage with male and female partners are linked to a high risk of HIV transmission and infection. Despite this, a large proportion of MSM throughout Asia and the Pacific lack access to HIV prevention and care programs [128, 129].

In the subgroup analyses, inconsistency between studies was alleviated. The highest prevalence of HIV (11.17%) was found in the studies sampling through Venue-day time sampling (VDTS), Syphilis (16.18%) in the studies through multiple methods, and co-infection (6.46%) in the studies used the Internet and venue-based sampling (Table 3). An almost opposite finding was observed in a previous study in China [50]. In England [130] individuals with high risk were willing to be tested with the VCT clinic or MSM network method. Therefore, a more appropriate and scientific sampling method needs to be applied in Asian regions as well for a more appropriate estimate of HIV, Syphilis, and their co-infection.

A few limitations of this study should be mentioned. Most of the selected studies were cross-sectional. Non-English literature has been excluded. For some regions or countries, no study was found. Therefore, findings from this meta-analysis and systematic review would not represent all regions of Asia. A large number of included studies used RDS, probability sampling, snowball sampling rather than population-based sampling which may lead the selection bias [50]. Another drawback is that the publication bias could not be avoided completely.

## 5 Conclusion

The findings of this systematic review will have several important implications. First, it will suggest a reappraisal of priorities and systems for HIV/STI surveillance in Asia to enable more robust, valid, and reliable prevalence estimates. Second, information from routine and periodic surveillance could be further validated and strengthened by highly focused, policy-relevant research to address key knowledge gaps in the epidemiological and socio-behavioral profile of specific sub-populations. The findings indicate that HIV, Syphilis, and their co-infection among MSM in Asia pacific region have increased markedly. To limit the spread of HIV and Syphilis infection, MSM should be targeted for increased HIV and Syphilis screening, and the development of efficient public health intervention programs is an important recommendation.

## Supporting information

PRISMA Checklist

MOOSE Checklist

## Data Availability

All data produced in the present work are contained in the manuscript

## 6 Declarations

### Ethics approval and consent to participate

This study is a systematic review and meta-analysis that used extracted data from the different published studies. Therefore, ethical approval and consent to participate are not relevant.

### Consent for publication

Not relevant for this study

### Availability of data and materials

All data produced in the present work are contained in the manuscript.

### Authors’ contributions

SM: Conceptualization, Formal analysis, Investigation, Methodology, Project administration, Software, Supervision, Validation, Visualization, Writing – original draft, Writing – review & editing

MM: Conceptualization, Formal analysis, Investigation, Methodology, Project administration, Software, Supervision, Validation, Visualization, Writing – original draft, Writing – review & editing

AM: Conceptualization, Data curation, Formal analysis, Investigation, Methodology, Project administration, Software, Supervision, Validation, Visualization, Writing – original draft, Writing – review & editing

SM: Data search, Formal analysis, Visualization, Writing – original draft, Writing – review & editing

MI: Data search, Formal analysis, Visualization, Writing – original draft, Writing – review & editing

AI: Data curation, Formal analysis, Visualization, Writing – original draft, Writing – review & editing

## Acknowledgements

None

## References

1. Anderson JE, Kanters S: Lack of sexual minorities’ rights as a barrier to HIV prevention among men who have sex with men and transgender women in Asia: a systematic review. LGBT health 2015, 2(1):16–26.

2. van Wijngaarden JWdL, Brown T, Girault P, Sarkar S, van Griensven F: The epidemiology of human immunodeficiency virus infection, sexually transmitted infections, and associated risk behaviors among men who have sex with men in the Mekong Subregion and China: implications for policy and programming. Sexually transmitted diseases 2009, 36(5):319–324.

3. Pham QD, Nguyen TV, Nguyen PD, Tran AT, Nguyen LT, Wilson DP, Zhang L: Men who have sex with men in southern Vietnam report high levels of substance use and sexual risk behaviours but underutilise HIV testing services: a cross-sectional study. Sex Transm Infect 2015, 91(3):178–182.

4. Morineau G, Nugrahini N, Riono P, Girault P, Mustikawati DE, Magnani R: Sexual risk taking, STI and HIV prevalence among men who have sex with men in six Indonesian cities. AIDS and Behavior 2011, 15(5):1033–1044.

5. Ekouevi DK, Bitty-Anderson AM, Gbeasor-Komlanvi FA, Salou M, Dagnra CA: Low prevalence of syphilis infection among key populations in Togo in 2017: a national cross-sectional survey. Archives of Public Health 2019, 77(1):1–6.

6. HIV/AIDS JUNPo: UNAIDS Data, Geneva, Switzerland; 2018. North American, Western and Central Europe: AIDS epidemic update regional summary 2019:1–16.

7. Narayanan P, Das A, Morineau G, Prabhakar P, Deshpande GR, Gangakhedkar R, Risbud A: An exploration of elevated HIV and STI risk among male sex workers from India. BMC public health 2013, 13(1):1059.

8. van Griensven F, Varangrat A, Wimonsate W, Tanpradech S, Kladsawad K, Chemnasiri T, Suksripanich O, Phanuphak P, MappStats PM, Kanggarnrua K: Trends in HIV prevalence, estimated HIV incidence, and risk behavior among men who have sex with men in Bangkok, Thailand, 2003–2007. JAIDS Journal of Acquired Immune Deficiency Syndromes 2010, 53(2):234–239.

9. van Griensven F, Thienkrua W, McNicholl J, Wimonsate W, Chaikummao S, Chonwattana W, Varangrat A, Sirivongrangson P, Mock PA, Akarasewi P: Evidence of an explosive epidemic of HIV infection in a cohort of men who have sex with men in Thailand. Aids 2013, 27(5):825–832.

10. Song D, Zhang H, Wang J, Liu Q, Wang X, Operario D, She M, Wang M, Zaller N: Prevalence and correlates of HIV infection and unrecognized HIV status among men who have sex with men and women in Chengdu and Guangzhou, China. AIDS and Behavior 2013, 17(7):2395–2404.

11. Dai W, Luo Z, Xu R, Zhao G, Tu D, Yang L, Wang F, Cai Y, Lan L, Hong F: Prevalence of HIV and syphilis co-infection and associated factors among non-commercial men who have sex with men attending a sexually transmitted disease clinic in Shenzhen, China. BMC infectious diseases 2017, 17(1):86.

12. Wang QQ, Chen XS, Yin YP, Liang GJ, Zhang RL, Jiang N, Huan XP, Yang B, Liu Q, Zhou YJ: HIV prevalence, incidence and risk behaviours among men who have sex with men in Yangzhou and Guangzhou, China: a cohort study. Journal of the International AIDS Society 2014, 17(1):18849.

13. Zhang Y, Chen P, Lu R, Liu L, Wu Y, Liu X, Zhao Z, Yi D: Prevalence of HIV among men who have sex with men in Chongqing, China, 2006–2009: cross–sectional biological and behavioural surveys. Sex Transm Infect 2012, 88(6):444–450.

14. Li R, Pan X, Ma Q, Wang H, He L, Jiang T, Wang D, Zhang Y, Zhang X, Xia S: Prevalence of prior HIV testing and associated factors among MSM in Zhejiang Province, China: a cross-sectional study. BMC public health 2016, 16(1):1152.

15. Mun M, Sopheab H, Tuot S, Morgan P, Pal K, Chhoun P: National HIV sentinel survey among women attending antenatal care clinics in Cambodia in 2014. Phnom Penh: National Center for HIV/AIDS. Dermatology and STD (NCHADS) 2016.

16. National AIDS Programme Department of Health MoH: Results of HIV Sentinel Sero-surveillance 2015, Myanmar. 2019.

17. Van Griensven F, Van Wijngaarden JWDL, Baral S, Grulich A: The global epidemic of HIV infection among men who have sex with men. Current Opinion in HIV and AIDS 2009, 4(4):300–307.

18. Dokubo EK, Kim AA, L. L-V, Nadol PJ, Prybylski D, Wolfe MI: HIV incidence in Asia: a review of available data and assessment of the epidemic. AIDS Rev 2013, 15(2):67–76.

19. van Griensven F, van Wijngaarden JWdL: A review of the epidemiology of HIV infection and prevention responses among MSM in Asia. Aids 2010, 24:S30–S40.

20. Ahn JY, Boettiger D, Kiertiburanakul S, Merati TP, Huy BV, Wong WW, Ditangco R, Lee MP, Oka S, Durier N: Incidence of syphilis seroconversion among HIV□infected persons in Asia: results from the TREAT Asia HIV Observational Database. Journal of the International AIDS Society 2016, 19(1):20965.

21. Mustanski BS, Newcomb ME, Du Bois SN, Garcia SC, Grov C: HIV in young men who have sex with men: a review of epidemiology, risk and protective factors, and interventions. Journal of sex research 2011, 48(2-3):218–253.

22. Young RM, Meyer IH: The trouble with “MSM” and “WSW”: Erasure of the sexual-minority person in public health discourse. American journal of public health 2005, 95(7):1144–1149.

23. Beyrer C: Hidden yet happening: the epidemics of sexually transmitted infections and HIV among men who have sex with men in developing countries. Sexually transmitted infections 2008, 84(6):410–412.

24. UNAIDS H: Men Who Have Sex with Men in Asia and the Pacific. UNAIDS, Geneva, Switzerland 2006.

25. Organization WH: HIV/AIDS in the South-East Asia Region, 2009. 2009.

26. Bowring A, Veronese V, Doyle J, Stoove M, Hellard M: HIV and sexual risk among men who have sex with men and women in Asia: a systematic review and meta-analysis. AIDS and Behavior 2016, 20(10):2243–2265.

27. Control CfD, Prevention: Outbreak of syphilis among men who have sex with men--Southern California, 2000. MMWR Morbidity and mortality weekly report 2001, 50(7):117.

28. Dilley J, Klausner JD, McFarland W, Kellogg T, Kohn R, Wong W, Louie B, Taylor MM, Kerndt PR, Carlos J: Trends in Primary and Secondary Syphilis and HIV Infections in Men Who Have Sex with Men---San Francisco and Los Angeles, California, 1998–2002. MMWR Morbidity and mortality weekly report 2004, 53(26):575.

29. Couturier E, Michel A, Janier M, Dupin N, Semaille C: Syphilis surveillance in France, 2000-2003. Euro surveillance: bulletin Europeen sur les maladies transmissibles= European communicable disease bulletin 2004, 9(12):8–10.

30. Dougan S, Evans BG, Elford J: Sexually transmitted infections in Western Europe among HIV-positive men who have sex with men. Sexually transmitted diseases 2007, 34(10):783–790.

31. Branger J, van der Meer JT, van Ketel RJ, Jurriaans S, Prins JM: High incidence of asymptomatic syphilis in HIV-infected MSM justifies routine screening. Sexually transmitted diseases 2009, 36(2):84–85.

32. Jin F, Prestage GP, Zablotska I, Rawstorne P, Imrie J, Kippax SC, Donovan B, Templeton DJ, Kaldor JM, Grulich AE: High incidence of syphilis in HIV-positive homosexual men: data from two community-based cohort studies. Sexual Health 2009, 6(4):281–284.

33. Ivens D, Patel M: Incidence and presentation of early syphilis diagnosed in HIV-positive gay men attending a central London outpatients’ department. International journal of STD & AIDS 2005, 16(3):201–202.

34. Feng Y, Wu Z, Detels R, Qin G, Liu L, Wang X, Wang J, Zhang L: HIV/STD prevalence among MSM in Chengdu, China and associated risk factors for HIV infection. Journal of acquired immune deficiency syndromes (1999) 2010, 53(Suppl 1):S74.

35. Choi K-H, Ning Z, Gregorich SE, Pan Q-c: The influence of social and sexual networks in the spread of HIV and syphilis among men who have sex with men in Shanghai, China. JAIDS Journal of Acquired Immune Deficiency Syndromes 2007, 45(1):77–84.

36. Sun M, Li D, Jin W, Jiang J, Guan L: Investigation on the Infection of HIV, HCV, Syphilis and HBV among MSM in Dalian City in 2008. Preventive Medicine Tribune 2009, 15(11):1074–1075.

37. Zhong F, Lin P, Xu H, Wang Y, Wang M, He Q, Fan L, Li Y, Wen F, Liang Y: Possible increase in HIV and syphilis prevalence among men who have sex with men in Guangzhou, China: results from a respondent-driven sampling survey. AIDS and Behavior 2011, 15(5):1058–1066.

38. Marcus U, Kollan C, Bremer V, Hamouda O: Relation between the HIV and the re-emerging syphilis epidemic among MSM in Germany: an analysis based on anonymous surveillance data. Sexually transmitted infections 2005, 81(6):456–457.

39. Pisani E, Girault P, Gultom M, Sukartini N, Kumalawati J, Jazan S, Donegan E: HIV, syphilis infection, and sexual practices among transgenders, male sex workers, and other men who have sex with men in Jakarta, Indonesia. Sexually transmitted infections 2004, 80(6):536–540.

40. Solomon SS, Srikrishnan AK, Sifakis F, Mehta SH, Vasudevan CK, Balakrishnan P, Mayer KH, Solomon S, Celentano DD: The emerging HIV epidemic among men who have sex with men in Tamil Nadu, India: geographic diffusion and bisexual concurrency. AIDS and Behavior 2010, 14(5):1001–1010.

41. Bozicevic I, Rode OD, Lepej SZ, Johnston LG, Stulhofer A, Dominkovic Z, Bacak V, Lukas D, Begovac J: Prevalence of sexually transmitted infections among men who have sex with men in Zagreb, Croatia. AIDS and Behavior 2009, 13(2):303–309.

42. Read P, Fairley, C.K. and Chow, E.P. : Increasing trends of syphilis among men who have sex with men in high income countries. Sexual health 2015, 12(2):155–163.

43. Vilayleck M: Continuing resurgence of syphilis in France. Weekly releases (1997–2007) 2001, 5(37):1686.

44. Botham SJ, Ressler K-A, Maywood P, Hope KG, Bourne CP, Conaty SJ, Ferson MJ, Mayne DJ: Men who have sex with men, infectious syphilis and HIV coinfection in inner Sydney: results of enhanced surveillance. Sexual health 2013, 10(4):291–298.

45. Hottes TS, Lindegger M, Consolacion T, Wong S, Lester RG, Montgomery CK, Doupe G, Morshed M, Ogilvie G, Gilbert M: Infectious Syphilis Among Gay, Bisexual and Other Men who Have Sex with Men in British Columbia: 2003 to 2012: Clinical Prevention Services Division, BC Centre for Disease Control; 2013.

46. Tan JY, Huedo-Medina TB, Warren MR, Carey MP, Johnson BT: A meta-analysis of the efficacy of HIV/AIDS prevention interventions in Asia, 1995–2009. Social science & medicine 2012, 75(4):676–687.

47. Wirtz AL, Kirey A, Peryskina A, Houdart F, Beyrer C: Uncovering the epidemic of HIV among men who have sex with men in Central Asia. Drug and alcohol dependence 2013, 132:S17–S24.

48. Owring AL, Veronese, V., Doyle, J.S., Stoove, M. and Hellard, M.: HIV and sexual risk among men who have sex with men and women in Asia: a systematic review and meta-analysis. AIDS and Behavior 2016, 20(10):2243–2265.

49. Chow EP, Wilson DP, Zhang L: HIV and syphilis co-infection increasing among men who have sex with men in China: a systematic review and meta-analysis. PloS one 2011, 6(8).

50. Gao L, Zhang L, Jin Q: Meta-analysis: prevalence of HIV infection and syphilis among MSM in China. Sexually transmitted infections 2009, 85(5):354–358.

51. Zhou Y, Li D, Lu D, Ruan Y, Qi X, Gao G: Prevalence of HIV and syphilis infection among men who have sex with men in China: a meta-analysis. BioMed research international 2014, 2014.

52. Chen G, Cao Y, Yao Y, Li M, Tang W, Li J, Babu GR, Jia Y, Huan X, Xu G: Syphilis incidence among men who have sex with men in China: results from a meta-analysis. International journal of STD & AIDS 2017, 28(2):170–178.

53. Mahmud S, Hossain S, Muyeed A, Islam MM, Mohsin M: The global prevalence of depression, anxiety, stress, and, insomnia and its’ changes among health professionals during COVID-19 pandemic: a rapid systematic review and meta-analysis. Heliyon 2021:e07393.

54. Liberati A, Altman DG, Tetzlaff J, Mulrow C, Gøtzsche PC, Ioannidis JP, Clarke M, Devereaux PJ, Kleijnen J, Moher D: The PRISMA statement for reporting systematic reviews and meta-analyses of studies that evaluate health care interventions: explanation and elaboration. Journal of clinical epidemiology 2009, 62(10):e1–e34.

55. Mahmud S, Mohsin M, Hossain S, Islam MM, Muyeed A: The Acceptance of COVID-19 Vaccine: A Global Rapid Systematic Review and Meta-Analysis. Available at SSRN 3855987 2021.

56. Mahmud S, Mohsin M, Khan IA, Mian AU, Zaman MA: Knowledge, beliefs, attitudes and perceived risk about COVID-19 vaccine and determinants of COVID-19 vaccine acceptance in Bangladesh. PloS one 2021, 16(9):e0257096.

57. Mahmud S, Mohsin M, Dewan MN, Muyeed A: The global prevalence of depression, anxiety, stress and insomnia among general population during COVID-19 pandemic: A systematic review and meta-analysis. 2021.

58. Guo Y, Li X, Song Y, Liu Y: Bisexual behavior among Chinese young migrant men who have sex with men: implications for HIV prevention and intervention. AIDS care 2012, 24(4):451–458.

59. Zou H, Wu Z, Yu J, Li M, Ablimit M, Li F, Pang L, Juniper N: Sexual risk behaviors and HIV infection among men who have sex with men who use the internet in Beijing and Urumqi, China. Journal of acquired immune deficiency syndromes (1999) 2010, 53(Suppl 1):S81.

60. Solomon SS, Mehta SH, Srikrishnan AK, Vasudevan CK, Mcfall AM, Balakrishnan P, Anand S, Nandagopal P, Ogburn E, Laeyendecker O: High HIV prevalence and incidence among men who have sex with men (MSM) across 12 cities in India. AIDS (London, England) 2015, 29(6):723.

61. Iedcr CJB, Icddr B: NATIONAL HIV SEROLOGICAL SURVEILLANCE, 2011 BANGLADESH.

62. Zhang Q, Deng P-j, Geng Y-j: Study on HIV/syphilis infections among men who have sex with men and their behavioral feature. China Tropical Medicine 2012, 12(2):219–220.

63. Wu Z, Xu J, Liu E, Mao Y, Xiao Y, Sun X, Liu Y, Jiang Y, McGoogan JM, Dou Z: HIV and syphilis prevalence among men who have sex with men: a cross-sectional survey of 61 cities in China. Clinical infectious diseases 2013, 57(2):298–309.

64. Feng L, Ding X, Lu R, Liu J, Sy A, Ouyang L, Pan C, Yi H, Liu H, Xu J: High HIV prevalence detected in 2006 and 2007 among men who have sex with men in China’s largest municipality: an alarming epidemic in Chongqing, China. JAIDS Journal of Acquired Immune Deficiency Syndromes 2009, 52(1):79–85.

65. Pham QD, Nguyen TV, Hoang CQ, Cao V, Van Khuu N, Phan HTT, Mai AH, Tran HN, Wilson DP, Zhang L: Prevalence of HIV/STIs and associated factors among men who have sex with men in An Giang, Vietnam. Sexually transmitted diseases 2012, 39(10):799–806.

66. Ministry of Health GoV: National Institute of Epidemiology and Family Health International. Results from HIV/STI integrated behavioral and biological surveillance (IBBS) in Vietnam, 2005–2006. Hanoi, Vietnam: Ministry of Health, Government of Vietnam 2007.

67. Huang S-W, Wang S-F, Cowó ÁE, Chen M, Lin Y-T, Hung C-P, Chen Y-H, Yang J-Y, Tang H-J, Chen Y-MA: Molecular epidemiology of HIV-1 infection among men who have sex with men in Taiwan in 2012. PLoS One 2015, 10(6):e0128266.

68. Kaphle Krdkks: Integrated Biological and Behavioral Surveillance (IBBS) Survey among Men who have Sex with Men (MSM) and Transgender (TG) People in Kathmandu Valley, Nepal. Round IV - 2012. 2013.

69. Bureau-Philippines E: Integrated HIV Behavioral and Serologic Surveillance: Philippines (Fact Sheets). 2013.

70. Control MoHNCfAaS, Teku K: Integrated Biological and Behavioral Surveillance (IBBS) Survey among Men who have Sex with Men (MSM) and Transgender (TG) in Kathmandu Valley. 2017.

71. Myanmar MoH: HIV Sentinel Sero□surveillance Survey Report. National AIDS Programme, Department of Health, Ministry of Health Myanmar 2012.

72. Leste DRoT: Results from the HIV/STI Integrated Biologic & Behavioral Surveillance (IBBS) Survey. 2011.

73. Zhao J, Cai W-D, Gan Y-X, Zhang Y, Yang Z-R, Cheng J-Q, Lin S-H, He M-L, Chen L, Wang X-R: A comparison of HIV infection and related risk factors between money boys and noncommercial men who have sex with men in Shenzhen, China. Sexually transmitted diseases 2012, 39(12):942–948.

74. Ruan S, Yang H, Zhu Y, Wang M, Ma Y, Zhao J, McFarland W, Raymond HF: Rising HIV prevalence among married and unmarried among men who have sex with men: Jinan, China. AIDS and Behavior 2009, 13(4):671–676.

75. Zhang L, Xiao Y, Lu R, Wu G, Ding X, Qian H-z, McFarland W, Ruan Y, Vermund SH, Shao Y: Predictors of HIV testing among men who have sex with men in a large Chinese city. Sexually transmitted diseases 2013, 40(3):235.

76. Fan S, Lu H, Ma X, Sun Y, He X, Li C, Raymond H, McFarland W, Sun J, Ma W: Behavioral and serologic survey of men who have sex with men in Beijing, China: implication for HIV intervention. AIDS patient care and STDs 2012, 26(3):148–155.

77. He Q, Wang Y, Lin P, Raymond HF, Li Y, Yang F, Zhao J, Li J, Ling L, McFarland W: High prevalence of risk behaviour concurrent with links to other high-risk populations: a potentially explosive HIV epidemic among men who have sex with men in Guangzhou, China. Sexually transmitted infections 2009, 85(5):383–390.

78. Ruan Y, Li D, Li X, Qian H-z, Shi W, Zhang X, Yang Z, Zhang X, Wang C, Liu Y: Relationship between syphilis and HIV infections among men who have sex with men in Beijing, China. Sexually transmitted diseases 2007, 34(8):592–597.

79. She M, Zhang H, Wang J, Xu J, Zhang Z, Fan Y, Sun Y, Zhang C: Associated factors for HIV and syphilis infection among men who have sex with men only and men who have sex with both men and women in cities of China. International journal of STD & AIDS 2013, 24(4):293–300.

80. Xiao Y, Sun J, Li C, Lu F, Allen KL, Vermund SH, Jia Y: Prevalence and correlates of HIV and syphilis infections among men who have sex with men in seven provinces in China with historically low HIV prevalence. JAIDS Journal of Acquired Immune Deficiency Syndromes 2010, 53:S66–S73.

81. Zeng Y, Zhang L, Li T, Lai W, Jia Y, Aliyu MH, Do M, Wang X, Han D, Huang W: Risk factors for HIV/syphilis infection and male circumcision practices and preferences among men who have sex with men in China. BioMed research international 2014, 2014.

82. Zhang L, Ding X, Lu R, Feng L, Li X, Xiao Y, Ruan Y, Vermund SH, Shao Y, Qian H-Z: Predictors of HIV and syphilis among men who have sex with men in a Chinese metropolitan city: comparison of risks among students and non-students. PLoS One 2012, 7(5):e37211.

83. Zhang X, Yu J, Li M, Sun X, Han Q, Li M, Zhou F, Li X, Yang Y, Xiao D: Prevalence and related risk behaviors of HIV, syphilis, and anal HPV infection among men who have sex with men from Beijing, China. AIDS and Behavior 2013, 17(3):1129–1136.

84. Runhua Li XP, Qiaoqin Ma, Hui Wang, Lin He, Tingting Jiang, Dayong Wang, Yan Zhang, Xingliang Zhang & Shichang Xia: Prevalence of prior HIV testing and associated factors among MSM in Zhejiang Province, China: a cross-sectional study. BMC Public Health 2016.

85. Lin Qu WW, Yongming Gao, Jingyuan Yang, Jijiang Dai, Dawei Wang & Bo Tao: A Cross-sectional Survey of HIV Transmission and Behavior among Men Who Have Sex with Men in Different Areas of Inner Mongolia Autonomous Region, China. BMC Public Health 2016.

86. Show less Xiping Huan M, Chun Hao, PhD, Hongjing Yan, M., Wenhui Guan, M., Xiaoqin Xu, M., Haitao Yang, M., Na Wang, M., Min Zhang, M., Weimin Tang, M., Jing Gu, PhD, Joseph T. F. Lau, PhD: High Prevalence of HIV and Syphilis Among Men Who Have Sex With Men Recruited by Respondent-Driven Sampling in a City in Eastern China. 2013.

87. Wenzhe Ma GW, Hui Zheng, Wenjuan Zhang, Zhihang Peng, Rongbin Yu, & Ning Wang: Prevalence and risk factors of HIV and syphilis, and knowledge and risk behaviors related to HIV/AIDS among men who have sex with men in Chongqing, China. J Biomed Res 2016.

88. Das A, Li J, Zhong F, Ouyang L, Mahapatra T, Tang W, Fu G, Zhao J, Detels R: Factors associated with HIV and syphilis co-infection among men who have sex with men in seven Chinese cities. International Journal of STD & AIDS 2015, 26(3):145–155.

89. Luo Y, Zhu C, Chen S, Geng Q, Fu R, Li X, Xu K, Cheng J, Ding J: Risk factors for HIV and syphilis infection among male sex workers who have sex with men: a cross-sectional study in Hangzhou, China, 2011. BMJ Open 2015, 5(4):e006791.

90. Yu Liu YR, Shiela M. Strauss, Lu Yin, Hongjie Liu, K. Rivet Amico, Chen Zhang, Yiming Shao, Han-Zhu Qian, Sten H. Vermund Dr.: Alcohol misuse, risky sexual behaviors, and HIV or syphilis infections among Chinese men who have sex with men. Drug and Alcohol Dependence 2016:Pages 239–246.

91. Wei S, Zhang, H., Wang, J., Song, D., Duan, Y., Yu, F., She, M., Wang, M. and Zhang, H.,: HIV and syphilis prevalence and associated factors among young men who have sex with men in 4 cities in China. AIDS and Behavior 2013:17(13), pp.1151–1158.

92. Zheng C, Xu, J.J., Hu, Q.H., Yu, Y.Q., Chu, Z.X., Zhang, J., Han, X.X., Lu, L., Wang, Z., Fu, J.H. and Chen, X., : Commercial sex and risk of HIV, syphilis, and herpes simplex virus-2 among men who have sex with men in six Chinese cities. BMC infectious diseases 2016:16(11), pp.11–11.

93. Gao W, Li, Z., Li, Y. and Qiao, X.: Sexual practices and the prevalence of HIV and syphilis among men who have sex with men in Lanzhou, China. Japanese journal of infectious diseases, 2015.

94. Wu J, Wu, H., Li, P. and Lu, C., : HIV/STIs risks between migrant MSM and local MSM: a cross-sectional comparison study in China. Peer J 2016:4, p.e2169.

95. Zhang X, Jia, M., Chen, M., Luo, H., Chen, H., Luo, W., Zhang, W., Ma, Y., Yang, C., Yang, Y. and Zhang, X.: Prevalence and the associated risk factors of HIV, STIs and HBV among men who have sex with men in Kunming, China.. International journal of STD & AIDS 2017:28(11), pp.1115–1123.

96. Shen L, Liu, X., Fu, G., Hao, S., Zhang, M., Wang, T., Yang, J., Wu, X. and Mao, L.: The epidemic of human immunodeficiency virus, hepatitis C virus, and syphilis infection, and the correlates of sexually transmitted infections among men who have sex with men in Zhenjiang, Jiangsu, China. Japanese journal of infectious diseases 2017.

97. Guanghua L, Yi, C., Shuai, T., Zhiyong, S., Zhenzhu, T., Yuhua, R., Yousuf, M.A. and Wensheng, F.: HIV, syphilis and behavioral risk factors among men who have sex with men in a drug-using area of southwestern China: Results of 3 cross-sectional surveys from 2013 to 2015. Medicine 2018.

98. Liu Y, Vermund, S.H., Ruan, Y., Liu, H., Zhang, C., Yin, L., Shao, Y. and Qian, H.Z.: HIV testing and sexual risks among migrant men who have sex with men: findings from a large cross-sectional study in Beijing, China.. AIDS care 2018:30(31), pp.86–94.

99. Liu YY, Tao, H.D., Liu, J., Fan, Y.G., Zhang, C., Li, P., Li, L.J., Huang, Q., Zhao, W. and Ye, D.Q.: Prevalence and associated factors of HIV infection among men who have sex with men in Hefei, China, 2013–2014: cross-sectional study. International journal of STD & AIDS 2016:27(24), pp.305–312.

100. Yang HT, Tang, W., Xiao, Z.P., Jiang, N., Mahapatra, T., Huan, X.P., Yin, Y.P., Wang, X.L., Chen, X.S. and Fu, G.F.: Worsening epidemic of HIV and syphilis among men who have sex with men in Jiangsu Province, China. Clinical Infectious Diseases 2014:58(12), pp.1753–1759.

101. Storm M, Deuba, K., Damas, J., Shrestha, U., Rawal, B., Bhattarai, R. and Marrone, G.: Prevalence of HIV, syphilis, and assessment of the social and structural determinants of sexual risk behaviour and health service utilisation among MSM and transgender women in Terai highway districts of Nepal: findings based on an integrated biological and behavioural surveillance survey using respondent driven sampling. BMC infectious diseases 2020: 20, pp.21–14.

102. Narayanan P, Das, A., Morineau, G., Prabhakar, P., Deshpande, G.R., Gangakhedkar, R. and Risbud, A.: An exploration of elevated HIV and STI risk among male sex workers from India. BMC public health 2013:13(11), pp.11–19.

103. Hussain T, Kulshreshtha, K.K. and Yadav, V.S.: Prevalence of HIV and syphilis infections among men who have sex with men attending an integrated counseling and testing centre at Agra: Comparison with studies in other regions of India. Journal of HIV/AIDS & Social Services 2018: 17(14), pp.353–368.

104. Kumta S, Lurie, M., Weitzen, S., Jerajani, H., Gogate, A., Row-kavi, A., Anand, V., Makadon, H. and Mayer, K.H.: Bisexuality, sexual risk taking, and HIV prevalence among men who have sex with men accessing voluntary counseling and testing services in Mumbai, India. Journal of acquired immune deficiency syndromes 2010: 53(52), p.227.

105. Cai WD, Zhao, J., Zhao, J.K., Raymond, H.F., Feng, Y.J., Liu, J., McFarland, W., Gan, Y.X., Yang, Z.R., Zhang, Y. and Tan, J.G.: HIV prevalence and related risk factors among male sex workers in Shenzhen, China: results from a time–location sampling survey. Sexually transmitted infections 2010:86(81), pp.15–20.

106. Xu JJ, Reilly, K.H., Lu, C.M., Ma, N., Zhang, M., Chu, Z.X., Wang, J.J., Yun, K. and Shang, H.: A cross-sectional study of HIV and syphilis infections among male students who have sex with men (MSM) in northeast China: implications for implementing HIV screening and intervention programs.. BMC Public Health 2011:11(11), pp.11–18.

107. Wang B, Li, X., Stanton, B., Liu, Y., & Jiang, S.: Socio-demographic and behavioral correlates for HIV and syphilis infections among migrant men who have sex with men in Beijing, China. AIDS Care:25(22), 249–257.

108. Project A: Integrated Biological and Behavioral Surveillance Survey (IBBS) among Men who have Sex with Men (MSM)) in the Kathmandu Valley Round III Family Health International /Nepal Baluwatar, Kathmandu, Nepal 2009.

109. International FH: Family Health International HIV/AIDS Prevention and Control Program Kathmandu, Nepal 2005 Integrated Bio-behavioral Survey (IBBS) among MSM Population in Kathmandu Valley, Final report HIV/AIDS Prevention and Control Program Kathmandu, Nepal

110. Health Do: National Epidemiology Center (2009) Integrated HIV Behavioral and Serologic Surveillance 2005-2009, Philippines (Fract sheet). 2009.

111. National STD/AIDS Control Programme MFPLaK: Integrated Biological and Behavioural Surveillance (IBBS) Survey among Key Populations at Higher Risk of HIV in Sri Lanka, 2014 – Report, Colombo,. 2015.

112. HIV/AIDS NCf: Dermatology and STDs (NCHADS) 2015 National HIV sentinel survey among ANC and MSM in Cambodia 2014.

113. Indonesia MoHRo: Integrated BiologicalBehavioral Surveillance 2011. 2011.

114. WHO NAPMa: HIV Sentinel Sero-Surveillance Survey 2008. 2009.

115. Colby D, Nguyen, N.A., Le, B. et al.: HIV and Syphilis Prevalence Among Transgender Women in Ho Chi Minh City, Vietnam.. AIDS Behav 2016:20, 379–385.

116. Prabawanti C, Bollen, L., Palupy, R. et al.: HIV, Sexually Transmitted Infections, and Sexual Risk Behavior Among Transgenders in Indonesia.. AIDS Behav 2011:15, 663–673.

117. Subramanian T, Ramakrishnan, L., Aridoss, S., Goswami, P., Kanguswami, B., Shajan, M., … Paranjape, R. S. : Increasing condom use and declining STI prevalence in high-risk MSM and TGs: evaluation of a largescale prevention program in Tamil Nadu, India. BMC Public Health 2013:13, 857.

118. Liu S, Zhao, J., Rou, K. et al.: A Survey of Condom Use Behaviors and HIV/STI Prevalence Among Venue-Based Money Boys in Shenzhen, China. AIDS Behav 2012:16, 835–846

119. Global AIDS monitoring 2019: indicators for monitoring the 2016 United Nations Political Declaration on HIV and AIDS. Geneva: UNAIDS; 2017.

120. Tsuboi M, Evans J, Davies EP, Rowley J, Korenromp EL, Clayton T, Taylor MM, Mabey D, Chico RM: Prevalence of syphilis among men who have sex with men: a global systematic review and meta-analysis from 2000–20. The Lancet Global Health 2021.

121. Chow EP, Wilson DP, Zhang L: HIV and syphilis co-infection increasing among men who have sex with men in China: a systematic review and meta-analysis. PloS one 2011, 6(8):e22768.

122. Yun K, Xu J, Reilly K, Zhang J, Jiang Y, Wang N, Shang H: Prevalence of bisexual behaviour among bridge population of men who have sex with men in China: a meta-analysis of observational studies. Sexually transmitted infections 2011, 87(7):563–570.

123. Wu MY, Gong HZ, Hu KR, Zheng H-y, Wan X, Li J: Effect of syphilis infection on HIV acquisition: a systematic review and meta-analysis. Sexually Transmitted Infections 2020.

124. UNAIDS: Global report: UNAIDS report on the global AIDS Epidemic 2019. 2020.

125. Rodrigo C, Rajapakse S: Current status of HIV/AIDS in South Asia. Journal of global infectious diseases 2009, 1(2):93.

126. Nepal B: Population mobility and spread of HIV across the Indo-Nepal border. Journal of health, population, and nutrition 2007, 25(3):267.

127. Organization WH: HIV/AIDS among men who have sex with men and transgender populations in South-East Asia: the current situation and national responses. 2010.

128. Avert: Global information and education on HIV and AIDS. 2020.

129. Pendse R, Gupta S, Yu D, Sarkar S: HIV/AIDS in the South-East Asia region: progress and challenges. Journal of virus eradication 2016, 2:1–6.

130. Sadler KE, McGarrigle CA, Elam G, Ssanyu-Sseruma W, Davidson O, Nichols T, Mercey D, Parry JV, Fenton KA: Sexual behaviour and HIV infection in black-Africans in England: results from the Mayisha II survey of sexual attitudes and lifestyles. Sexually transmitted infections 2007, 83(7):523–529.

